# Disordered eating and self-harm as risk factors for poorer mental health during the COVID-19 pandemic: A UK-based birth cohort study

**DOI:** 10.1101/2021.04.30.21256377

**Authors:** Naomi Warne, Jon Heron, Becky Mars, Alex S. F. Kwong, Francesca Solmi, Rebecca Pearson, Paul Moran, Helen Bould

**Affiliations:** Centre for Academic Mental Health, Population Health Sciences, Bristol Medical School, University of Bristol, UK; Population Health Sciences, Bristol Medical School, University of Bristol, UK; MRC Integrative Epidemiology Unit, Bristol Medical School, University of Bristol, UK; Division of Psychiatry, Centre for Clinical Brain Sciences, University of Edinburgh, UK; UCL Division of Psychiatry, UK; Gloucestershire Health and Care NHS Foundation Trust, Gloucester, UK

**Author notes:** **<u>Corresponding author:</u>** Dr Naomi Warne, Centre for Academic Mental Health, Population Health Sciences, University of Bristol, Oakfield House, Oakfield Grove, Bristol, BS8 2BN.

**Keywords:** ALSPAC, COVID-19, disordered eating, self-harm, mental health, pandemic, lockdown

## Abstract

**Background:** Young adults and especially those with pre-existing mental health conditions, such as disordered eating and self-harm, appear to be at greater risk of developing metal health problems during the COVID-19 pandemic. However, it is unclear whether this increased risk is affected by any changes in lockdown restrictions, and whether any lifestyle changes could moderate this increased risk.

**Methods:** In a longitudinal UK-based birth cohort (The Avon Longitudinal Study of Parents and Children, ALSPAC) we assessed the relationship between pre-pandemic measures of disordered eating and self-harm and mental health during the COVID-19 pandemic in 2,657 young adults. Regression models examined the relationship between self-reported disordered eating, self-harm, and both disordered eating and self-harm at age 25 years and depressive symptoms, anxiety symptoms and mental wellbeing during a period of eased restrictions in the COVID-19 pandemic (May-July 2020) when participants were aged 27-29 years. Analyses were adjusted for sex, questionnaire completion date, pre-pandemic socioeconomic disadvantage and pre-pandemic mental health and wellbeing. We also examined whether lifestyle changes (sleep, exercise, alcohol, visiting green space, eating, talking with family/friends, hobbies, relaxation) in the initial UK lockdown (April-May 2020) moderated these associations.

**Results:** Pre-existing disordered eating, self-harm and comorbid disordered eating and self-harm were all associated with the reporting of a higher frequency of depressive symptoms and anxiety symptoms, and poorer mental wellbeing during the pandemic compared to individuals without disordered eating and self-harm. Associations remained when adjusting for pre-pandemic mental health measures. There was little evidence that interactions between disordered eating and self-harm exposures and lifestyle change moderators affected pandemic mental health and wellbeing.

**Conclusions:** Young adults with pre-pandemic disordered eating, self-harm and comorbid disordered eating and self-harm were at increased risk for developing symptoms of depression, anxiety and poor mental wellbeing during the COVID-19 pandemic, even when accounting for pre-pandemic mental health. Lifestyle changes during the pandemic do not appear to alter this risk. A greater focus on rapid and responsive service provision is essential to reduce the impact of the pandemic on the mental health of these already vulnerable individuals.

**Plain English summary:** The aim of this project was to explore the mental health of young adults with disordered eating behaviours (such as fasting, vomiting/taking laxatives, binge-eating and excessive exercise) and self-harm during the COVID-19 pandemic. We analysed data from an established study that has followed children from birth (in 1991 and 1992) up to present day, including during the pandemic when participants were 28 years old. We looked at the relationship between disordered eating and/or self-harm behaviours from *before* the pandemic and mental health problems (symptoms of depression and anxiety) and mental wellbeing *during* the pandemic. We also explored whether there were any lifestyle changes (such as changes in sleep, exercise, visiting green space) that might be linked to better mental health and wellbeing in young adults with disordered eating and self-harm. We found that young adults with prior disordered eating and/or self-harm had more symptoms of depression and anxiety, and worse mental wellbeing than individuals without prior disordered eating or self-harm. However, lifestyle changes did not appear to affect mental health and wellbeing in these young adults. Our findings suggest that people with a history of disordered eating and/or self-harm were at high risk for developing mental health problems during the pandemic, and they will need help from mental health services.

## Background

The coronavirus disease 2019 (COVID-19) pandemic has had an immense impact on people’s lives worldwide. On 23^rd^ March 2020, a national lockdown was announced as a UK public health strategy instructing the public to stay at home except for certain limited purposes. During this time, people could only leave their homes once a day for one hour for exercise or shopping for essential goods (e.g. food, medicine). Non-essential businesses were closed, schools were closed for the majority of students (exceptions were for vulnerable children and children of keyworkers), and people were urged to work from home where possible. Restrictions were gradually eased, allowing for unlimited exercise outside (13^th^ May 2020), groups of up to six people to meet outside with social distancing (1^st^ June 2020), year groups returning to schools (1^st^ June 2020), and non-essential shops re-opening (15^th^ June 2020). The pandemic and associated restrictions radically changed people’s lives and there is evidence that mental health in the UK population was worse during lockdown than before the pandemic (1–4). Furthermore, young adults and individuals with prior mental health problems were at increased risk of common mental health problems (depressive symptoms and anxiety symptoms) during the UK lockdown (1).

Young adults with pre-existing disordered eating and self-harm are likely to be at particularly high risk of experiencing poor mental health during the pandemic. Eating disorder behaviours and self-harm are common in young adults (5) and are associated with increased risk of mortality and psychiatric comorbidity (6–11), which may be exacerbated by circumstances in the pandemic. Disordered eating and self-harm are common (12,13) and commonly co-occur in clinical (14) and general population (5) samples. This comorbidity is a great clinical concern as it increases the risk of poorer overall mental health (15–17) and risk for suicide (18), compared with when the behaviours occur in isolation. Nevertheless, eating disorders and self-harm remain highly under-researched (19,20) and more work is required to ensure that we understand the needs of people with eating disorders and/or self-harm during the pandemic (21,22). There is growing concern that individuals with a history of eating disorders and/or self-harm may have been particularly affected by the pandemic and the associated restrictions. In addition to the broader risk factors that can affect many people (for instance social isolation, stressful life events) individuals with eating disorders may have experienced more specific risk factors potentially leading to increased distress such as changes in access to food, exercise limitations, media messaging and restricted healthcare access (21,23). Individuals with disordered eating are also likely to experience some of these more specific risk factors. Similarly, the pandemic has resulted in a number of exacerbating factors for self-harm (22) and the reductions of clinical services presentations for self-harm may reflect reduced help-seeking in this vulnerable group (24,25). Given these additional risk factors, individuals with eating disorders/disordered eating and self-harm may be a key ‘at risk’ groups for poorer mental health (such as depression and anxiety) during the pandemic. Such common mental health problems are important to research and detect early as they are associated with significant impairment (26), as well as poor health, education and social outcomes (27–31). Furthermore, depression and anxiety are common concerns for young people with eating disorders during the pandemic, with 73% of individuals reporting the pandemic has increased their feelings of depression and 77% reporting the pandemic has increased their feelings of anxiety (32). Prior research has found that disordered eating and self-harm were risk factors for increased depressive and anxiety symptoms in young adults during the first UK lockdown (1). However, it is unclear whether this increased risk persists when lockdown restrictions have eased, and whether lifestyle factors could play a role in this risk.

To date, there has been limited published research on COVID-19-related common mental health outcomes in individuals with eating disorders and self-harm. One cross-sectional Australian study found that individuals self-reporting an eating disorder had higher levels of depression, anxiety and stress than individuals not reporting an eating disorder in April 2020 (33). Cross-sectional studies have found the majority of participants with eating disorders have also self-reported a worsening of depressive symptoms (32,34) and anxiety symptoms (32,35,36) due to the pandemic, however a longitudinal study found no change in general psychopathology (including depression and anxiety) pre-to post-lockdown in eating disorder patients (37). There is very limited research for pandemic common mental health problems in those with a history of self-harm but there is emerging evidence that individuals with a previous history of self-harm with suicidal intent (suicide attempt) were more likely to have depression in a Greek lockdown (April-May 2020) than those without a history of suicide attempt (38).

Although informative, conclusions from these studies are limited by the self-selected participants, the small sample sizes (typically under ∼200 people) and the retrospective reporting of changes in mental health. Furthermore, many studies focus on clinical samples, whereas only a minority of individuals with disordered eating and self-harm seek help (6,8,39). Research using birth cohorts with pre-pandemic measures of mental health can provide a more accurate representation of effects on mental health coincident with the pandemic in these groups. Such samples include a more representative sample of participants than those found from convenience sampling and include participants with disordered eating or self-harm who may be missed by clinical services. As the COVID-19 pandemic and mitigation efforts have been a universal exposure it is difficult to disentangle effects of the pandemic from natural changes in risk over time. Nevertheless, longitudinal studies with pre-pandemic information allow for adjustment for potential confounding factors and prior mental health problems so we can be more confident that associations are coincident with the pandemic rather than due to confounding factors or prior mental health problems. In this study, we used a UK-based birth cohort 1) to investigate whether prior disordered eating and self-harm were risk factors for higher levels of depression and anxiety and lower levels of mental wellbeing during a period of eased restrictions in the COVID-19 pandemic; and 2) to assess whether lifestyle changes can identify individuals with prior disordered eating and self-harm who may have better mental health in the pandemic. The first of these is important to help inform who may be at greater risk of mental health problems during the current pandemic and for informing who may require ongoing help from mental health services. The second is important to provide insights into factors that may help improve mental health in the current pandemic or pandemics to come.

## Methods

### Sample

We analysed data on young adults from a UK-based birth cohort: The Avon Longitudinal Study of Parents and Children (ALSPAC) (40–42). Pregnant women living in the Avon area of Bristol (UK) with an expected delivery date between 1^st^ April 1991 and 31^st^ December 1992 were invited to take part in the study. 14,541 pregnant women were enrolled in ALSPAC and had completed at least one assessment (questionnaire or in person clinic) by 19^th^ July 1999. This resulted in 14,062 live births, and 13,988 children who were alive at 1 year of age. We used data on offspring from this core sample only. Ethical approval for the study was obtained from the ALSPAC Ethics and Law Committee and the Local Research Ethics Committees.

Data were collected at multiple time points and from multiple informants via regular questionnaires and face-to-face assessments at research clinics. Some of these data were collected and managed using REDCap electronic data capture tools (43,44). Please note that the study website contains details of all the data that is available through a fully searchable data dictionary and variable search tool: http://www.bristol.ac.uk/alspac/researchers/our-data/. In this study, we focused on exposures of disordered eating and self-harm behaviours reported by participants *before* the pandemic at age 25 years, and additional data on other health characteristics, collated from two online questionnaires sent following UK lockdown (which started on 23rd March 2020). Mental health and wellbeing outcomes were self-reported during a period of eased restrictions (26th May - 4th July 2020) in the COVID-19 pandemic when participants were age 28 years (45). We also looked at self-reported lifestyle changes during the initial UK lockdown (9th April - 15th May 2020) (46) as potential moderators of this relationship (see timeline of assessments in Figure 1).

**Figure 1.**
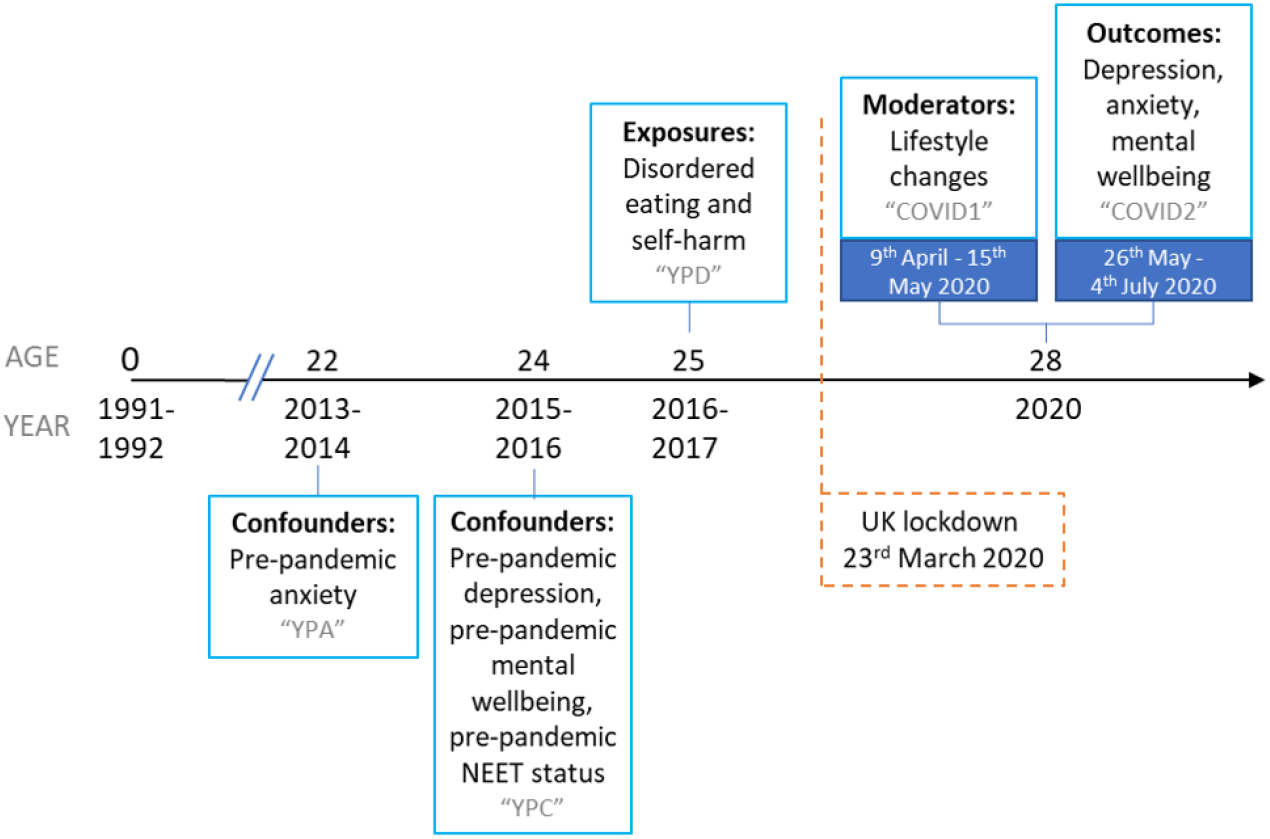
Timeline of ALSPAC assessments NEET = Not in education, employment or training. Grey text indicates the name of the ALSPAC questionnaire.

We conducted our primary analyses on an imputed sample of 2657 individuals (1891 (71.17%) females, 766 (28.83%) males) who completed questions on lifestyle changes during the pandemic (see Supplementary Figure 1 for flowchart of attrition). Responders to this survey were more likely than non-responders to be female, white, in education, employment or training prior to the pandemic, and have a higher maternal education and have parents who owned a home around birth (see Supplementary Table 1).

### Exposures

Disordered eating and self-harm were self-reported via questionnaire at age 25 (“YPD” questionnaire). Age at completion ranged from 23.8 years to 26.3 years with a mean age of 24.8 (SD 0.5) years.

#### Exposure 1: Disordered eating

Disordered eating was measured using the Youth Risk Behaviour Surveillance System questionnaire (47). We used questions about behaviours in the last year to lose weight or avoid gaining weight: 1) fasting for at least a day; 2) purging (vomiting or taking laxatives/other medications); 3) excessive exercise (exercise that frequently interfered with daily routine/work, or frequently exercising even when sick/injured); as well as 4) binge-eating with a sense of loss of control. Our primary exposure of interest was *any disordered eating*, a composite measure derived for any report of fasting, purging, excessive exercise or binge-eating at any frequency. Secondary exposures of interest were each of the individual disordered eating behaviours (fasting, purging, binge-eating, excessive exercise) at any frequency, and a composite measure of any of these behaviours at least once a week in line with DSM-5 diagnostic criteria (48): *DSM-5 frequency disordered eating*. This variable was derived based on frequency of disordered eating alone. We did not make a diagnosis of an eating disorder and did not incorporate any other diagnostic criteria into the DSM-5 frequency disordered eating variable. Questions, possible responses and variable derivation are presented in Supplementary Table 2.

#### Exposure 2: Self-harm

We assessed self-harm using questions adapted from the Child and Adolescent Self-Harm in Europe study (49). We used self-harm behaviour in the last year, in order to be comparable with our measures of disordered eating. Participants were asked a series of questions about the presence and frequency of self-harm behaviours (see Supplementary Table 2). Our primary exposure of interest was *any self-harm*, regardless of suicidal intent, in the past year. This was derived from questions asking whether participants had hurt themselves on purpose in any way and how many times they did this in the last year. Secondary exposures of interest were *self-harm without suicidal intent* and *self-harm with suicidal intent (suicidal attempt)*. Self-harm without suicidal intent was reported if, when asked the question “when was the last time you hurt yourself on purpose, without intending to kill yourself?”, they responded with “in the last week”, or “more than a week ago but in the last year”. Similarly, participants with those responses to the question “when was the last time you hurt yourself on purpose and you seriously wanted to kill yourself?” were recorded as having *self-harm with suicidal intent*. Participants could therefore report both self-harm *with* suicidal intent and self-harm *without* suicidal intent in the last year.

#### Exposure 3: Comorbid disordered eating and self-harm

Individuals who reported *any disordered eating* (at any frequency) and *any self-harm* were coded as having comorbid disordered eating and self-harm behaviours.

### Outcomes

Outcomes were taken from the second COVID-19-related questionnaire (“COVID2” questionnaire) which was sent to participants during a period of eased restrictions between 26th May and 4th July 2020. Participants were aged 27-29 years at completion (mean (SD) = 28.2 (0.5) years).

#### Outcome 1: Depressive symptoms during the COVID-19 pandemic

Depressive symptoms were measured using the short version of the Mood and Feelings Questionnaire (sMFQ) (50). Participants reported whether 13 depressive symptom statements were ‘not true’ (0), ‘sometimes true’ (1), and ‘true’ (2) for the previous 2 weeks. Scores were summed (possible range 0-26) with higher scores indicating more depressive symptoms. As all scales used are sum-scored comprised of a set of ordinal responses we followed the approach described by Flora (2020) (51) to derive coefficient omega as a measure of reliability. We used the lavaan package (52) in R and the “WLSMV” estimator so reliability is derived from polychoric rather than Pearson correlations. Reliability of the sMFQ was excellent (omega = 0.923).

#### Outcome 2: Anxiety symptoms during the COVID-19 pandemic

Anxiety symptoms were measured on the Generalised Anxiety Disorder 7-item questionnaire (GAD-7) (53). Participants reported whether they had been bothered by each of the 7 anxiety statements in the past 2 weeks on the response scale ‘not at all’ (0), ‘several days’ (1), ‘more than half the days’ (2), and ‘nearly every day’ (3). These responses were summed for a total score with possible range 0-21, with higher scores indicating higher levels of anxiety. Reliability of the GAD-7 was excellent (omega = 0.933).

#### Outcome 3: Mental wellbeing during the COVID-19 pandemic

Mental wellbeing was measured on the Warwick-Edinburgh Mental Well-Being Scale (WEMWBS) (54). This consisted of 14 wellbeing statements that participants rated over the past 2 weeks on a scale of ‘none of the time’ (1), ‘rarely’ (2), ‘some of the time’ (3), ‘often’ (4), and ‘all of the time’ (5). Summed scores produced a possible range of 14-70 with lower scores indicating poorer mental wellbeing. Reliability of the WEMWBS was excellent (omega = 0.927).

### Moderating factors

Participants reported on a number of lifestyle changes that occurred after the first UK lockdown was announced (23^rd^ March 2020) in the first online COVID-19-related questionnaire (“COVID1” questionnaire) (10). This questionnaire was sent during lockdown between 9th April and 15th May 2020 (mean (SD) age = 28.1 (0.5) years). Questions about lifestyle changes included changes in the amount of sleep, exercise, alcohol drunk, visiting green space, practising relaxation/mindfulness/mediation, eating, as well as changes in time spent talking to family/friends outside their home and time spent doing hobbies/things they enjoy. Participants reported whether the amount they did each activity had “decreased”, “stayed the same”, “increased” or was “not applicable” since the first UK lockdown (23^rd^ March 2020). For our analyses we combined “not applicable” with “stayed the same” as we made the assumption that the majority of people responding “not applicable” likely maintained an absence of the activity.

### Confounders and descriptive variables

Based on their plausible associations with exposures, outcomes and moderating factors, we adjusted for hypothesised confounders of sex, completion date of the COVID1 questionnaire, pre-pandemic socioeconomic status and pre-pandemic mental health symptoms. Sex was recorded at birth by the fieldworkers who visited the maternity units. Participation in education, employment or training activities was used as the pre-pandemic indicator of socioeconomic status, with those not in education, employment, or training (NEET) designated a being in the socioeconomic disadvantage category. Participants reported on their education and employment status, prior to the pandemic (2015-2016) at age 24 years (see Supplementary Table 3 for question wording and variable coding). For pre-pandemic mental health symptoms, we used the most recent reports on the same questionnaires preceding the disordered eating and self-harm exposures (age 25). For depressive symptoms (sMFQ; omega = 0.921) and mental wellbeing (WEMWBS; omega = 0.930) this was at age 24 years and for anxiety symptoms (GAD-7; omega = 0.914) this was at age 22 years. Timings of all exposures, outcomes and confounders are presented in Figure 1.

We also described the sample’s sociodemographic characteristics, and pandemic-related experiences, the latter were measured on the COVID2 questionnaire. Participants were categorised as *living alone* if they responded “no I live on my own” to the question “Do you live with anyone?”. Participants were defined as a *keyworker* if they responded “yes” to the question: “Are you a keyworker, or has your work been classified as critical to the COVID-19 response?”. Participants were considered to be *furloughed during the pandemic* if they responded “yes” to the question: “Which of these would you say best describes your current situation now?: Employed but on paid leave (including furlough)”. For pandemic financial situation, participants were asked: “Overall, how do you feel your current financial situation compares to how it was before the COVID-19 pandemic?” with possible responses of “I’m much worse off”, “I’m a little worse off”, “I’m about the same”, “I’m a little better off”, “I’m much better off”. Individuals responding “I’m much worse off” and “I’m a little worse off” were coded as having *financial problems during the pandemic*. Full information on question wording and response options are provided in Supplementary Table 4.

### Analysis

All analyses were performed in Stata version 16 (55). First, we described the samples in terms of sex, race, sociodemographic factors, pandemic-related experiences, and key exposure and outcome variables. Secondly, we used linear regression models to explore the relationship between disordered eating and self-harm exposures and the three mental health and wellbeing outcomes (depressive symptoms, anxiety symptoms, mental wellbeing) during a period of eased restrictions in the COVID-19 pandemic. We focused on our primary exposures of *any disordered eating, any self-harm* and *comorbid disordered eating and self-harm*. Analyses were repeated with secondary exposures of specific types of disordered eating (fasting, purging, excessive exercise, binge eating, DSM-5 frequency disordered eating) and self-harm with and without suicidal intent. We conducted these analyses unadjusted for confounders (Model A) and with sequential adjustment to assess the effects of confounders. We progressively adjusted for: sex, COVID1 questionnaire completion date, pre-pandemic NEET status (Model B); and corresponding pre-pandemic mental health symptoms/wellbeing (Model C). We conducted sensitivity analyses to further examine the effect of disordered eating severity by repeating regression models using a disordered eating exposure variable coded for no disordered eating (0, reference category), less frequent disordered eating at less than once a week (1) and DSM-5 frequency disordered eating at least once a week in line with DSM-5 diagnostic criteria (2).

We examined whether the association between exposure and outcome varied depending on changes in lifestyle factors by testing whether there was evidence of an interaction between lifestyle change and disordered eating status, and lifestyle change and self-harm status on pandemic mental health and wellbeing. To aid in interpretation of interaction results we also examined the association between disordered eating and self-harm exposures and lifestyle change moderators, as well as associations between lifestyle change factors and pandemic-related mental health outcomes.

#### Missing data

Primary analyses were conducted on an imputed dataset of 2657 individuals with complete data on lifestyle change moderators measured on the COVID1 questionnaire. Missing data on primary exposures, outcomes, and confounders were imputed (see Supplementary Table 5 for amount of missing data). Secondary exposures were not imputed, and we did not impute data on lifestyle changes in COVID1 questionnaire or pandemic-related descriptive factors as these data were unique and unlikely to be explained by auxiliary data in ALSPAC. Data were imputed using the multivariate imputation by chained equations (MICE) approach (56) under the missing at random (MAR) assumption. In addition to variables used in the main analysis, we incorporated auxiliary variables related to the missing data mechanism. Auxiliary variables were disordered eating at 16 and 18 years, self-harm at 18 and 24 years, Body Mass Index at 18 and 24 years, and mental health and wellbeing measures from the COVID1 questionnaire. A number of sociodemographic variables collected near birth including maternal age, parity, maternal education, maternal social class, paternal social class, home ownership status and birthweight were also used. One hundred datasets were imputed (a decision informed by studying the Monte Carlo errors for the estimated parameters). Separate multiple imputation models were performed: one model for the main regression models and multiple imputation models including all interactions (57) to allow for interactions between each exposure and each lifestyle change variable. We compared estimates from imputed analysis to analysis of observed data with complete information on disordered eating, self-harm, mental health measures during the pandemic, lifestyle change moderators, and confounders (see flowchart of attrition, Supplementary Figure 1).

## Results

### Descriptive results

Descriptive information for the imputed and observed samples is presented in Table 1. The sample was predominantly white and before the pandemic the majority of participants were in education, employment or training (i.e. did not have NEET status). During the pandemic (observed sample), 7.20% were living alone, approximately two in five (39.66%) participants were keyworkers, one in eight (12.31%) had been furloughed, and a quarter (24.61%) reported financial problems. At age 25 (imputed sample), 32.04% of the sample reported some form of disordered eating in the past year, 8.97% reported self-harm in the past year, and 5.53% reported comorbid disordered eating and self-harm in the past year. The most common specific disordered eating behaviour (observed sample) was binge-eating (20.46%), whereas the most common type of self-harm reported was self-harm without suicidal intent (6.19%).

**Table 1.**
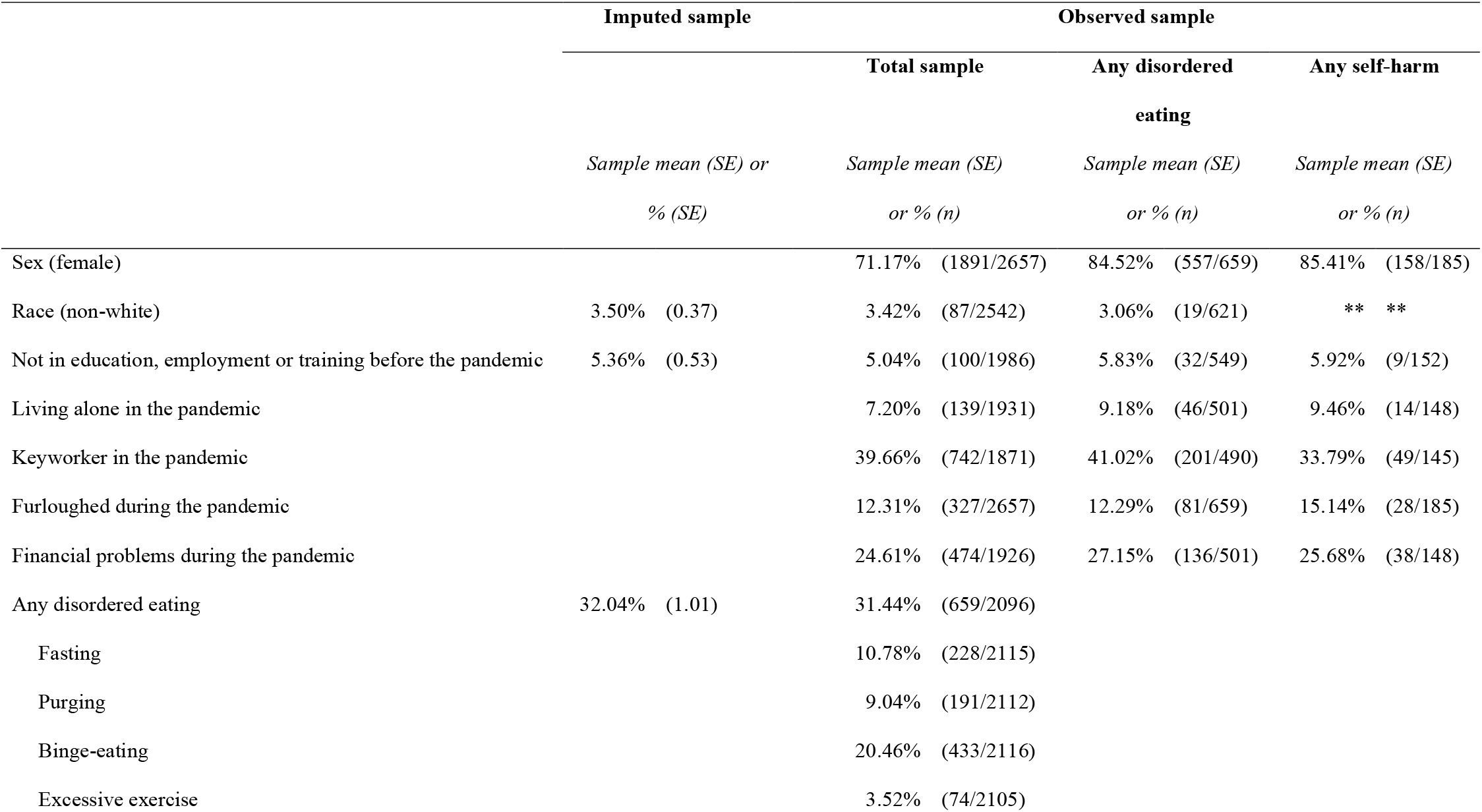
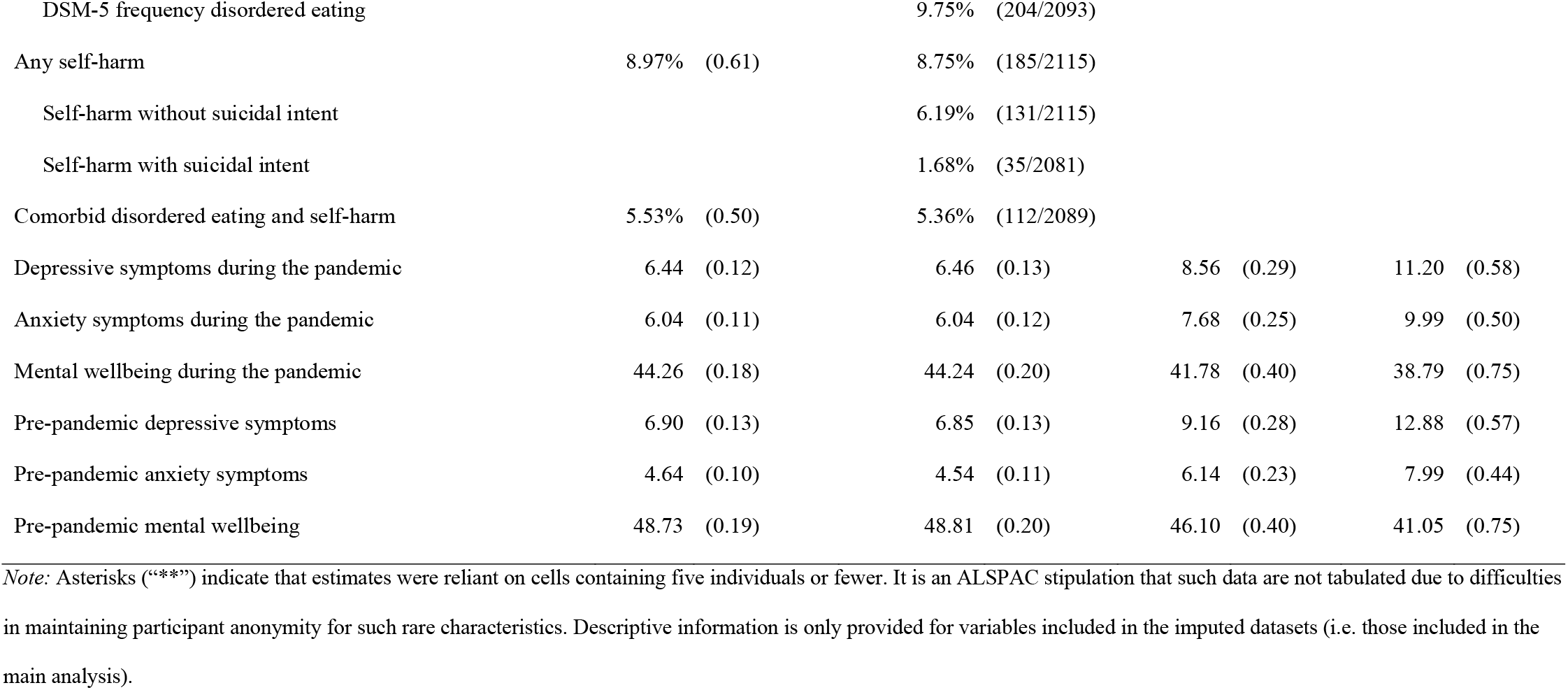
Descriptive information on imputed and observed samples

### Associations between disordered eating/self-harm and mental health outcomes during the pandemic

Regression models testing the associations between disordered eating and self-harm exposures with mental health and wellbeing outcomes are presented in Table 2. There was evidence that a history of any disordered eating (DE), any self-harm (SH), and comorbid disordered eating and self-harm (DE+SH) were associated with higher depressive and anxiety symptoms, and lower mental wellbeing during the pandemic. These associations were present in unadjusted models for depressive symptoms (B_DE_ (95% CI) = 2.98 (2.44, 3.53), p < .001; B_SH_ (95% CI) = 5.19 (4.31, 6.08), p < .001; B_DE+SH_ (95% CI) = 6.15 (5.02, 7.29), p < .001), anxiety symptoms (B_DE_ (95% CI) = 2.43 (1.92, 2.95), p < .001; B_SH_ (95% CI) = 4.55 (3.74, 5.36), p < .001; B_DE+SH_ (95% CI) = 5.38 (4.30, 6.47), p < .001), and mental wellbeing (B_DE_ (95% CI) = −3.49 (−4.30, −2.67), p < .001; B_SH_ (95% CI) = −5.78 (−7.06, −4.50), p < .001; B_DE+SH_ (95% CI) = −7.81 (−9.43, −6.19), p < .001). Associations remained unchanged when adjusting for sex, COVID questionnaire completion date and pre-pandemic NEET status for depressive symptoms (B_DE_ (95% CI) = 2.72 (2.17, 3.27), p < .001; B_SH_ (95% CI) = 4.95 (4.07, 5.83), p < .001; B_DE+SH_ (95% CI) = 5.87 (4.74, 7.00), p < .001), anxiety symptoms (B_DE_ (95% CI) = 2.11 (1.59, 2.63), p < .001; B_SH_ (95% CI) = 4.26 (3.46, 5.05), p < .001; B_DE+SH_ (95% CI) = 5.05 (3.98, 6.12), p < .001), and mental wellbeing (B_DE_ (95% CI) = −3.35 (−4.18, −2.51), p < .001; B_SH_ (95% CI) = −5.62 (−6.90, −4.33), p < .001; B_DE+SH_ (95% CI) = −7.60 (−9.21, −6.00), p < .001). There was still strong evidence of these associations after further adjustment for pre-pandemic mental health and wellbeing although the magnitude of the associations was attenuated for depressive symptoms (B_DE_ (95% CI) = 1.37 (0.84, 1.90), p < .001; B_SH_ (95% CI) = 2.13 (1.24, 3.01), p < .001; B_DE+SH_ (95% CI) = 2.52 (1.38, 3.66)), p < .001), anxiety symptoms (B_DE_ (95% CI) = 1.24 (0.74, 1.74), p < .001; B_SH_ (95% CI) = 2.69 (1.87, 3.50), p < .001; B_DE+SH_ (95% CI) = 3.08 (2.01, 4.15), p < .001), and mental wellbeing (B_DE_ (95% CI) = −1.82 (−2.59, −1.06), p < .001; B_SH_ (95% CI) = −2.18 (−3.43, −0.93), p < .001; B_DE+SH_ (95% CI) = − 3.64 (−5.19, −2.09), p < .001). These results suggest that disordered eating, self-harm, and comorbid disordered eating and self-harm were risk factors for mental health problems coincident with the pandemic. Results using imputed data were consistent with complete case observed data (see Supplementary Table 6).

**Table 2.** Association between disordered eating and self-harm exposures and pandemic mental health and wellbeing outcomes

| Exposure | Unadjusted Model A |  | Adjusted Model B |  | Fully Adjusted Model C |  |
| --- | --- | --- | --- | --- | --- | --- |
|  | B (95% CI) | p | B (95% CI) | p | B (95% CI) | p |
| ***Outcome: Depressive symptoms*** |  |  |  |  |  |  |
| Disordered eating | 2.98 (2.44, 3.53) | <.001 | 2.72 (2.17, 3.27) | <.001 | 1.37 (0.84, 1.90) | <.001 |
| Self-harm | 5.19 (4.31, 6.08) | <.001 | 4.95 (4.07, 5.83) | <.001 | 2.13 (1.24, 3.01) | <.001 |
| Comorbid disordered eating and self-harm | 6.15 (5.02, 7.29) | <.001 | 5.87 (4.74, 7.00) | <.001 | 2.52 (1.38, 3.66) | <.001 |
| ***Outcome: Anxiety symptoms*** |  |  |  |  |  |  |
| Disordered eating | 2.43 (1.92, 2.95) | <.001 | 2.11 (1.59, 2.63) | <.001 | 1.24 (0.74, 1.74) | <.001 |
| Self-harm | 4.55 (3.74, 5.36) | <.001 | 4.26 (3.46, 5.05) | <.001 | 2.69 (1.87, 3.50) | <.001 |
| Comorbid disordered eating and self-harm | 5.38 (4.30, 6.47) | <.001 | 5.05 (3.98, 6.12) | <.001 | 3.08 (2.01, 4.15) | <.001 |
| ***Outcome: Mental wellbeing*** |  |  |  |  |  |  |
| Disordered eating | -3.49 (-4.30, -2.67) | <.001 | -3.35 (-4.18, -2.51) | <.001 | -1.82 (-2.59, -1.06) | <.001 |
| Self-harm | -5.78 (-7.06, -4.50) | <.001 | -5.62 (-6.90, -4.33) | <.001 | -2.18 (-3.43, -0.93) | <.001 |
| Comorbid disordered eating and self-harm | -7.81 (-9.43, -6.19) | <.001 | -7.60 (-9.21, -6.00) | <.001 | -3.64 (-5.19, -2.09) | <.001 |

Results using imputed data (n=2657). Model A = unadjusted; Model B = adjusted for sex, COVID1 questionnaire completion date and pre-pandemic socioeconomic status; Model C = adjusted for sex, COVID questionnaire completion date, pre-pandemic socioeconomic status and pre-pandemic mental health and wellbeing measures.

Broadly similar patterns were detected for analyses with secondary exposures (see Supplementary Table 7). Subtypes of disordered eating (fasting, purging, binge-eating, DSM-5 frequency disordered eating) and self-harm (with and without suicidal intent) were associated with greater depressive symptoms, greater anxiety symptoms and lower mental wellbeing during the pandemic in unadjusted and in adjusted models. However, excessive exercise was not associated with pandemic depressive symptoms (B (95% CI) = 0.75 (−0.75, 2.25), p = 0.324) and mental wellbeing (B (95% CI) = −0.19 (−2.49, 2.12), p = 0.875) in fully adjusted models. In addition, self-harm without suicidal intent was no longer associated with mental wellbeing in the pandemic when accounting for pre-pandemic mental wellbeing (B (95% CI) = −1.28 (−2.93, 0.37), p = 0.127).

Frequency of disordered eating was important (see Supplementary Table 8). In all fully adjusted models, compared to no disordered eating, both less frequent disordered eating (less than once a week) and more frequent DSM-5 level disordered eating (once a week or more) were associated with worse mental health outcomes coincident with the pandemic. However, these associations were stronger for the more frequent DSM-5 level of symptoms. This was the case for depressive symptoms (B_less frequent_ (95% CI) = 1.17 (0.50, 1.83), p = .001; B_more frequent_ (95% CI) = 2.31 (1.35, 3.27), p < .001), anxiety symptoms (B_less frequent_ (95% CI) = 0.99 (0.33, 1.66), p = .003; B_more frequent_ (95% CI) = 1.97 (1.00, 2.95), p < .001) and mental wellbeing (B_less frequent_ (95% CI) = −1.54 (−2.54, −0.54), p = .002; B_more frequent_ (95% CI) = −2.31 (−3.77, −0.85), p = .002).

### Moderation of lifestyle changes

Interaction results are presented in Figure 2 alongside associations between disordered eating/self-harm exposures and pandemic mental health and wellbeing outcomes stratified by levels of lifestyle change moderators to aid interpretation (see Supplementary Table 9 for frequencies of each lifestyle change). As can be seen, irrespective of lifestyle change and outcome, disordered eating and self-harm were all associated with worse mental health outcomes, but there was little evidence of interaction effects between lifestyle changes during the pandemic and prior disordered eating (Figure 2a, 2c and 2e) and prior self-harm (Figure 2b, 2d and 2f). There was evidence for one interaction between disordered eating and sleep on anxiety symptoms (Figure 2c, F = 5.07, p = 0.006), in that keeping the same level of sleep was associated with lower levels of anxiety than changes (decreases and increases) in sleep for those with disordered eating. This pattern of results was also seen for depressive symptom and mental wellbeing outcomes but there was little evidence for interaction effects (depressive symptoms Figure 2a, F = 1.96, p =.142; mental wellbeing Figure 2e, F = 2.35, p = 0.096). There was an interaction between self-harm and hobbies for anxiety symptoms (Figure 2d, F = 3.10, p = 0.045), whereby changes (increases and decreases) in time spent on hobbies was associated with fewer anxiety symptoms for those with a history of self-harm. However, on the whole, the lack of strong evidence for associations suggests that lifestyle changes are unlikely to impact those with disordered eating or self-harm differently to those without these problems.

**Figure 2.**
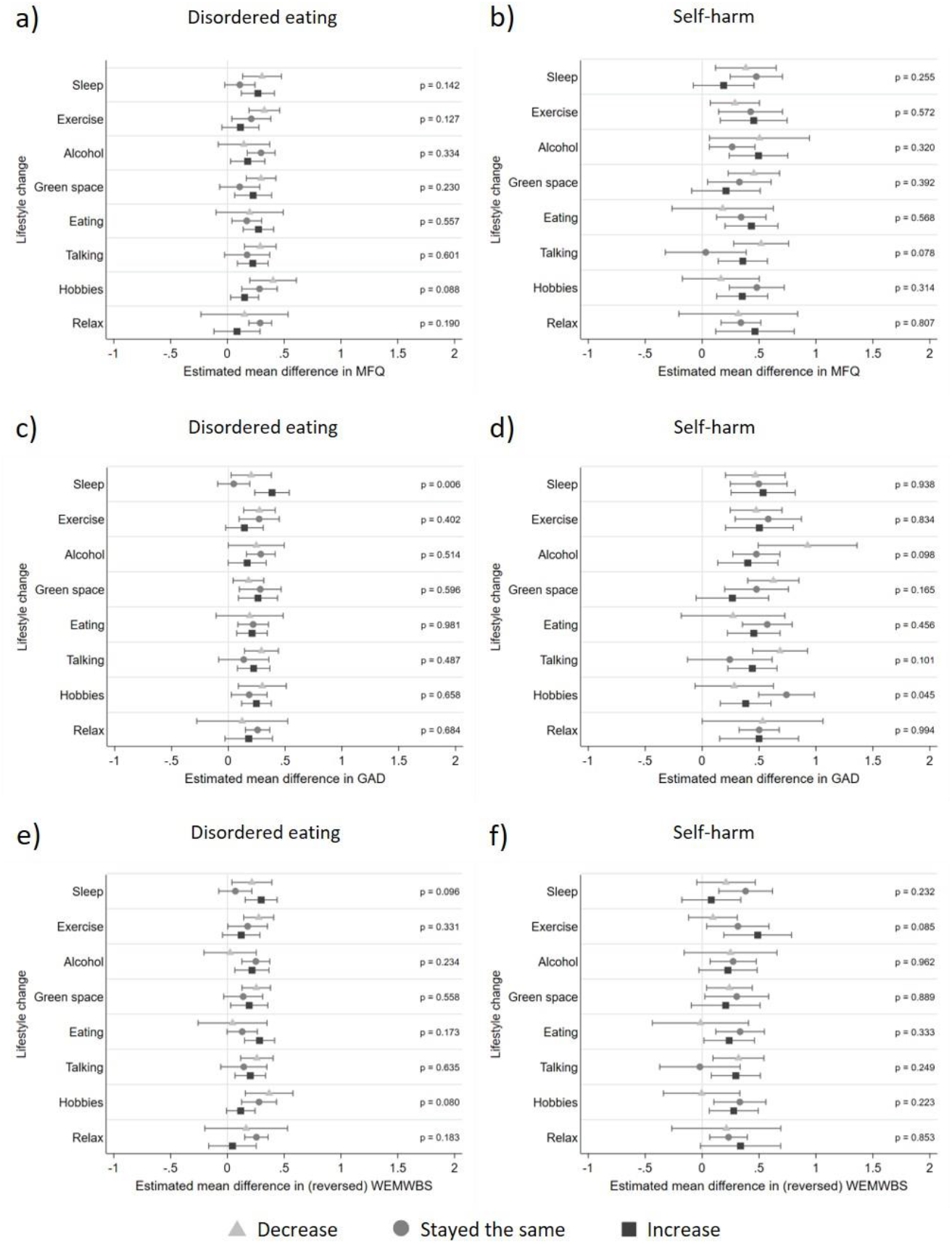
Adjusted associations between disordered eating/self-harm and mental health and wellbeing stratified by moderators (lifestyle changes) Panel a) Association between disordered eating and depressive symptoms, stratified by moderators; Panel b) Association between self-harm and depressive symptoms, stratified by moderators; Panel c) association between disordered eating and anxiety symptoms, stratified by moderators; Panel d) association between self-harm and anxiety symptoms, stratified by moderators; Panel e) association between disordered eating and mental wellbeing, stratified by moderators; Panel f) association between self-harm and mental wellbeing, stratified by moderators. All associations were adjusted for sex, date of completion for COVID1 questionnaire, pre-pandemic NEET (not in education, employment or training) status and pre-pandemic symptoms. Results using imputed data (n=2657). P values displayed are for the interactions between exposure and moderator on mental health and wellbeing outcomes. MFQ = Mood and Feelings Questionnaire for depressive symptoms GAD = Generalised Anxiety Disorder 7-item questionnaire for anxiety symptoms WEMWBS = Warwick-Edinburgh Mental Well-Being Scale for mental wellbeing

There was evidence for some associations between disordered eating and self-harm exposures and lifestyle changes (Supplementary Table 10). We also found that changes (decreases and increases) in lifestyle were generally associated with poorer subsequent mental health and wellbeing compared to maintaining the same level of each lifestyle factor (Supplementary Table 11). This suggests that any lifestyle changes during the pandemic (even those generally seen as positive such as increases in visiting green space and increases in time spent doing hobbies) were linked to worse subsequent mental health in the whole sample, regardless of prior history of disordered eating or self-harm. Overall these results highlight that lifestyle change moderators are not independent of exposures and outcomes.

## Discussion

In this UK-based birth cohort, young adults with a previous history of either disordered eating, self-harm, or comorbid disordered eating and self-harm, were at increased risk for poor mental health and wellbeing during the COVID-19 pandemic. Specifically, disordered eating, self-harm and comorbid disordered eating and self-harm were associated with higher depressive symptoms, higher anxiety symptoms and poorer mental wellbeing during a period of easing restrictions. Associations attenuated somewhat with the addition of prior mental health and wellbeing confounders, but there was still evidence of associations between disordered eating and self-harm exposures and pandemic mental health outcomes independent of pre-pandemic levels of mental health and wellbeing. Furthermore, there was limited evidence that any changes in lifestyle (sleep, exercise, drinking alcohol, visiting green space, eating, talking with family/friends outside the home, hobbies, relaxation techniques) during lockdown restrictions moderated these relationships. These findings suggest that young adults with disordered eating and self-harm may need additional help to prevent mental health problems from developing during the pandemic or for rapid access to treatment.

Previous research in this cohort has found individuals with a history of disordered eating (at DSM-5 frequency (48)) and self-harm were associated with higher levels of depressive and anxiety symptoms during the initial UK lockdown (1). The current study extends these findings to a period of eased restrictions and suggests that individuals with disordered eating and self-harm may be at enduring risk for common mental health problems throughout the COVID-19 pandemic. We also found that the association between a history of disordered eating behaviours and poorer mental health during the pandemic was not limited to more severe DSM-5 level frequencies but was also present for less frequent levels of disordered eating. These findings have implications for the current pandemic in addition to pandemics that may occur in the future. We suggest that policymakers and healthcare professionals should be made aware that individuals with disordered eating and self-harm are at increased risk of experiencing worse mental health outcomes during periods of lockdown and eased restrictions. Prioritising more funding for mental health services in the current pandemic and in advance of future pandemics would enable healthcare professionals to support and treat more individuals. Intervening as early as possible would help prevent negative effects of enduring mental health problems that coincide with periods of lockdown and eased restrictions.

We found little evidence that lifestyle changes could moderate the risk of prior disordered eating and self-harm on mental health outcomes during the pandemic. Given the numerous tests conducted we are cautious about over-interpreting weak evidence of moderation. However, we found evidence for an interaction between disordered eating and sleep on anxiety symptoms, whereby maintaining pre-pandemic levels of sleep was associated with fewer anxiety symptoms than increases or decreases in sleep for those with a history of disordered eating. This is difficult to interpret given lifestyle changes in the pandemic (in this case sleep) are unlikely to be independent of exposures and outcomes (see Supplementary Tables 10 and 11) and may be affected by concurrent or recent changes in mental health, employment, and childcare responsibilities. In general, changes in the lifestyle factors assessed were associated with worse mental health and wellbeing regardless of prior disordered eating or self-harm (Supplementary Table 11). Future work focusing on factors that promote resilience in these at risk groups, who are already at elevated risk of mortality (7,9), is warranted. It is not clear what would be useful for young adults with prior disordered eating and self-harm during the pandemic, but it is likely that access to mental health services will be important. This is difficult in the context of the pandemic when service use has reduced (58). Therefore, additional funding that enables greater service provision, which is adapted for pandemic-related restrictions, will likely be necessary.

The strengths of this study include the large sample size, the use of longitudinal data to assess mental health coincident with the pandemic by accounting for pre-pandemic mental health, and use of participants with impairing disordered eating and self-harm who would normally be omitted from clinical samples. Nevertheless, the study should be viewed in light of its limitations. First, we were unable to explore disordered eating and self-harm during the pandemic as there was no data available for this. We were therefore unable to explore whether increases in depressive symptoms, anxiety symptoms and poorer mental wellbeing could be driven by changes in disordered eating and self-harm behaviours. Second, we are unable to estimate a direct effect of the COVID-19 pandemic as it is a universal exposure and there is no control group. However, using ALSPAC has enabled us to control for pre-pandemic mental health and confounding factors in analyses so we are confident that associations found are coincident with the COVID-19 pandemic, and not a consequence of previous depression, anxiety or poor mental wellbeing. Third, we used imputed datasets under the assumption that data was missing at random, which if not true, could mean the results are biased. Furthermore, we only imputed up to those with complete lifestyle change data on the COVID1 questionnaire and this may limit generalisability of findings as this sample was more likely to be female, white and have less socioeconomic disadvantage than those who did not respond to the survey. Fourth, ALSPAC attempted to recruit all pregnant mothers due to give birth in the Avon area but those recruited were under-representative of minority ethnic groups living in the area at the time (ALSPAC cohort: 2.2% ethnic minority groups; Avon population at the time: 4.1% ethnic minority groups) and were also more affluent than the general Avon population (41). The majority of ALSPAC participants are white and we were unable to provide further breakdown of the “non-white” group, as due to small numbers, this data was censored to preserve anonymity. Therefore we were unable to assess the effect of ethnicity on the relationships between disordered eating, self-harm and pandemic mental health. This is important as individuals from different ethnic minority groups are known to have health inequalities, which have been exacerbated during the COVID-19 pandemic. Further research is vital to examine risk and potentially helpful factors in these groups, ideally in the context of population-based cohort samples which more accurately represent the ethnic and socioeconomic diversity of the UK. Fifth, ALSPAC collected sex at birth and did not collect gender at the timepoints in this study. This is important as rates of disordered eating and self-harm are likely to be higher in transgender adults and adolescents (59,60) and there is evidence that transgender adults might have had worse mental health outcomes as a result of the pandemic (61). Sixth, interpretation of our moderation results is difficult given lifestyle change moderators were not independent of the exposures and outcomes, and may be associated with additional factors such as employment change. Consequently, we were unable to make inferences about the direction of effect, and this required further exploration. Finally, we did not correct for multiple comparisons due to the exploratory nature of analyses and the lack of independence of many of the measures. These results are therefore in need of replication in independent samples.

## Conclusions

Individuals with prior disordered eating and self-harm were at increased risk of developing common mental health problems during the COVID-19 pandemic. There was little evidence to support lifestyle changes moderating this risk. Further work is needed to identify factors that might increase resilience among individuals with disordered eating and self-harm during the pandemic in order to prevent them from developing common mental health problems. Additional funding for mental health services will likely be important to provide rapid treatment for these at risk young adults.

## Data Availability

ALSPAC data access is through a system of managed open access. The steps below highlight how to apply for access to the data included in this paper and all other ALSPAC data. 1. Please read the ALSPAC access policy (http://www.bristol.ac.uk/media-library/sites/alspac/documents/researchers/data-access/ALSPAC_Access_Policy.pdf) which describes the process of accessing the data and samples in detail, and outlines the costs associated with doing so. 2. You may also find it useful to browse our fully searchable research proposals database, which lists all research projects that have been approved since April 2011. 3. Please submit your research proposal (https://proposals.epi.bristol.ac.uk/) for consideration by the ALSPAC Executive Committee. You will receive a response within 10 working days to advise you whether your proposal has been approved. If you have any questions about accessing data or samples, please (data) or (samples).

## List of abbreviations

ALSPAC: Avon Longitudinal Study of Parents and Children
GAD7: Generalised Anxiety Disorder 7-item questionnaire
MFQ: Mood and Feelings Questionnaire
WEMWBS: Warwick-Edinburgh Mental Well-Being Scale

## Declarations

### Ethics approval and consent to participate

Ethics approval for the study was obtained from the ALSPAC Ethics and Law Committee and the Local Research Ethics Committees.

### Consent for publication

Not applicable.

### Availability of data and materials

ALSPAC data access is through a system of managed open access. The steps below highlight how to apply for access to the data included in this paper and all other ALSPAC data.

1. Please read the ALSPAC access policy (PDF, 843kB) which describes the process of accessing the data and samples in detail, and outlines the costs associated with doing so.
2. You may also find it useful to browse our fully searchable research proposals database, which lists all research projects that have been approved since April 2011.
3. Please submit your research proposal for consideration by the ALSPAC Executive Committee. You will receive a response within 10 working days to advise you whether your proposal has been approved.

If you have any questions about accessing data or samples, please (data) or (samples).

## Authors*’* contributions

All authors were involved in conception and design of the study. NW and JH performed data analysis. NW wrote the first draft of the manuscript. All authors contributed to interpretation of results. All authors were involved in critically revising the manuscript for important intellectual content and approval of the final manuscript.

## Acknowledgments

We are extremely grateful to all the families who took part in this study, the midwives for their help in recruiting them, and the whole ALSPAC team, which includes interviewers, computer and laboratory technicians, clerical workers, research scientists, volunteers, managers, receptionists and nurses.

## References

1. Kwong ASF, Pearson RM, Adams MJ, Northstone K, Tilling K, Smith D, et al. Mental health before and during the COVID-19 pandemic in two longitudinal UK population cohorts. Br J Psychiatry. 2020;24:1–10.

2. Dickerson J, Kelly B, Lockyer B, Bridges S, Cartwright C, Willan K, et al. “When will this end? Will it end?” the impact of the March-June 2020 UK Covid-19 lockdown response on mental health: A longitudinal survey of mothers in the Born in Bradford study. medRxiv. 2020;2020.11.30.20239954.

3. Office for National Statistics. Coronavirus and depression in adults, Great Britain: June 2020 [Internet]. 2020 [cited 2021 Feb 18]. Available from: https://www.ons.gov.uk/peoplepopulationandcommunity/wellbeing/articles/coronavirusanddepressioninadultsgreatbritain/june2020

4. Pierce M, Hope H, Ford T, Hatch S, Hotopf M, John A, et al. Mental health before and during the COVID-19 pandemic: a longitudinal probability sample survey of the UK population. The Lancet Psychiatry. 2020;7(10):883–92.

5. Warne N, Heron J, Mars B, Moran P, Stewart A, Munafò M, et al. Comorbidity of self-harm and disordered eating in young people: Evidence from a UK population-based cohort. J Affect Disord. 2021;282:386–90.

6. Treasure J, Duarte TA, Schmidt U. Eating disorders. Lancet. 2020;395(10227):899–911.

7. Arcelus J, Mitchell AJ, Wales J, Nielsen S. Mortality rates in patients with anorexia nervosa and other eating disorders: A meta-analysis of 36 studies. Arch Gen Psychiatry. 2011 Jul;68(7):724–31.

8. Hawton K, Saunders KEA, O’Connor RC. Self-harm and suicide in adolescents. Lancet. 2012;379(9834):2373–82.

9. Skegg K. Self-harm. Lancet. 2005;366(9495):1471–83.

10. Carroll R, Metcalfe C, Gunnell D. Hospital Presenting Self-Harm and Risk of Fatal and Non-Fatal Repetition: Systematic Review and Meta-Analysis. Gluud LL, editor. PLoS One [Internet]. 2014 Feb 28 [cited 2020 Jul 22];9(2):e89944. Available from: https://dx.plos.org/10.1371/journal.pone.0089944

11. Mars B, Heron J, Crane C, Hawton K, Lewis G, Macleod J, et al. Clinical and social outcomes of adolescent self harm: population based birth cohort study. BMJ [Internet]. 2014 Oct 21 [cited 2021 Aug 25];349. Available from: https://www.bmj.com/content/349/bmj.g5954

12. Smink FRE, Van Hoeken D, Oldehinkel AJ, Hoek HW. Prevalence and severity of DSM-5 eating disorders in a community cohort of adolescents. Int J Eat Disord. 2014;47(6):610–9.

13. Gillies D, Christou MA, Dixon AC, Featherston OJ, Rapti I, Garcia-Anguita A, et al. Prevalence and Characteristics of Self-Harm in Adolescents: Meta-Analyses of Community-Based Studies 1990–2015. J Am Acad Child Adolesc Psychiatry [Internet]. 2018;57(10):733–41. Available from: https://doi.org/10.1016/j.jaac.2018.06.018

14. Svirko E, Hawton K. Self-Injurious Behavior and Eating Disorders: The Extent and Nature of the Association. Suicide Life-Threatening Behav. 2007;37(4):409–21.

15. Claes L, Jiménez-Murcia S, Agüera Z, Castro R, Sánchez I, Menchõn JM, et al. Male eating disorder patients with and without non-suicidal self-injury: A comparison of psychopathological and personality features. Eur Eat Disord Rev. 2012;20(4):335–8.

16. Claes L, Vandereycken W, Vertommen H. Eating-disordered patients with and without self-injurious behaviours: A comparison of psychopathological features. Eur Eat Disord Rev. 2003;11(5):379–96.

17. Wright F, Bewick BM, Barkham M, House AO, Hill AJ. Co-occurrence of self-reported disordered eating and self-harm in UK university students. Br J Clin Psychol. 2009;48(4):397–410.

18. Brausch AM, Perkins NM. Nonsuicidal self-injury and disordered eating: Differences in acquired capability and suicide attempt severity. Psychiatry Res. 2018;266:72–8.

19. Solmi F, Bould H, Lloyd EC, Lewis G. The shrouded visibility of eating disorders research. The Lancet Psychiatry [Internet]. 2021 Feb 1 [cited 2021 Aug 24];8(2):91–2. Available from: http://www.thelancet.com/article/S2215036620304235/fulltext

20. MQ. UK Mental Health Research Funding 2014-2017 [Internet]. 2019. Available from: https://www.mqmentalhealth.org/wp-content/uploads/UKMentalHealthResearchFunding2014-2017digital.pdf

21. Touyz S, Lacey H, Hay P. Eating disorders in the time of COVID-19. J Eat Disord. 2020;8(1):19.

22. Holmes EA, O’Connor RC, Perry VH, Tracey I, Wessely S, Arseneault L, et al. Multidisciplinary research priorities for the COVID-19 pandemic: a call for action for mental health science. The Lancet Psychiatry. 2020;7(6):547–60.

23. Cooper M, Reilly EE, Siegel JA, Coniglio K, Sadeh-Sharvit S, Pisetsky EM, et al. Eating disorders during the COVID-19 pandemic and quarantine: an overview of risks and recommendations for treatment and early intervention. Eat Disord. 2020;–23.

24. Kapur N, Clements C, Appleby L, Hawton K, Steeg S, Waters K, et al. Effects of the COVID-19 pandemic on self-harm. The Lancet Psychiatry. 2021;8(2):e4.

25. Hawton K, Casey D, Bale E, Brand F, Ness J, Waters K, et al. Self-harm during the early period of the COVID-19 pandemic in England: Comparative trend analysis of hospital presentations. J Affect Disord. 2021 Mar 1;282:991–5.

26. Kessler RC. The costs of depression. Psychiatr Cinics North Am. 2012 Mar;35(1):1–14.

27. Kawakami N, Abdulghani EA, Alonso J, Bromet EJ, Bruffaerts R, Caldas-de-Almeida JM, et al. Early-life mental disorders and adult household income in the World Mental Health Surveys. Biol Psychiatry. 2012 Aug;72(3):228–37.

28. Lee S, Tsang A, Breslau J, Aguilar-Gaxiola S, Angermeyer M, Borges G, et al. Mental disorders and termination of education in high-income and low-and middle-income countries: Epidemiological study. Br J psychiatry. 2009 May;194(5):411–7.

29. Kupferberg A, Bicks L, Hasler G. Social functioning in Major Depressive Disorder. Neurosci Biobehav Rev. 2016 Oct 1;69:313–32.

30. Davis L, Uezato A, Newell JM, Frazier E. Major depression and comorbid substance use disorders. Curr Opin Psychiatry. 2008;21(1):14–8.

31. Essau CA, Lewinsohn PM, Olaya B, Seeley JR. Anxiety disorders in adolescents and psychosocial outcomes at age 30. J Affect Disord. 2014 Jul 1;163:125–32.

32. Vitagliano JA, Jhe G, Milliren CE, Lin JA, Spigel R, Freizinger M, et al. COVID-19 and eating disorder and mental health concerns in patients with eating disorders. J Eat Disord 2021 91 [Internet]. 2021 Jul 2 [cited 2021 Aug 25];9(1):1–8. Available from: https://jeatdisord.biomedcentral.com/articles/10.1186/s40337-021-00437-1

33. Phillipou A, Meyer D, Neill E, Tan EJ, Toh WL, Van Rheenen TE, et al. Eating and exercise behaviors in eating disorders and the general population during the COVID-19 pandemic in Australia: Initial results from the COLLATE project. Int J Eat Disord. 2020 Jul 1;53(7):1158–65.

34. Schlegl S, Maier J, Meule A, Voderholzer U. Eating disorders in times of the COVID-19 pandemic—Results from an online survey of patients with anorexia nervosa. Int J Eat Disord. 2020;53(11):1791–800.

35. Termorshuizen JD, Watson HJ, Thornton LM, Borg S, Flatt RE, MacDermod CM, et al. Early impact of COVID-19 on individuals with self-reported eating disorders: A survey of ∼1,000 individuals in the United States and the Netherlands. Int J Eat Disord. 2020;53(11):1780–90.

36. Fernández-Aranda F, Casas M, Claes L, Bryan DC, Favaro A, Granero R, et al. COVID-19 and implications for eating disorders. Eur Eat Disord Rev. 2020;28(3):239–45.

37. Castellini G, Cassioli E, Rossi E, Innocenti M, Gironi V, Sanfilippo G, et al. The impact of COVID-19 epidemic on eating disorders: A longitudinal observation of pre versus post psychopathological features in a sample of patients with eating disorders and a group of healthy controls. Int J Eat Disord. 2020;53(11):1855–62.

38. Fountoulakis KN, Apostolidou MK, Atsiova MB, Filippidou AK, Florou AK, Gousiou DS, et al. Self-reported changes in anxiety, depression and suicidality during the COVID-19 lockdown in Greece. J Affect Disord. 2021;279:624–9.

39. Solmi F, Hotopf M, Hatch SL, Treasure J, Micali N. Eating disorders in a multi-ethnic inner-city UK sample: prevalence, comorbidity and service use. Soc Psychiatry Psychiatr Epidemiol. 2016;51(3):369–81.

40. Boyd A, Golding J, Macleod J, Lawlor DA, Fraser A, Henderson J, et al. Cohort profile: The ‘Children of the 90s’-The index offspring of the Avon Longitudinal Study of Parents and Children. Int J Epidemiol. 2013;42(1):111–27.

41. Fraser A, Macdonald-wallis C, Tilling K, Boyd A, Golding J, Davey smith G, et al. Cohort profile: The avon longitudinal study of parents and children: ALSPAC mothers cohort. Int J Epidemiol. 2013;42(1):97–110.

42. Northstone K, Lewcock M, Groom A, Boyd A, Macleod J, Timpson N, et al. The Avon Longitudinal Study of Parents and Children (ALSPAC): an update on the enrolled sample of index children in 2019. Wellcome Open Res. 2019;4:51.

43. Harris PA, Taylor R, Minor BL, Elliott V, Fernandez M, O’Neal L, et al. The REDCap consortium: Building an international community of software platform partners. J Biomed Inform. 2019;95:103208.

44. Harris PA, Taylor R, Thielke R, Payne J, Gonzalez N, Conde JG. Research electronic data capture (REDCap)-A metadata-driven methodology and workflow process for providing translational research informatics support. J Biomed Inform. 2009;42(2):377–81.

45. Northstone K, Smith D, Bowring C, Wells N, Crawford M, Haworth S, et al. The Avon Longitudinal Study of Parents and Children - A resource for COVID-19 research: Questionnaire data capture May-July 2020. Wellcome Open Res. 2020;5:210.

46. Northstone K, Howarth S, Smith D, Bowring C, Wells N, Timpson NJ. The Avon Longitudinal Study of Parents and Children - A resource for COVID-19 research: Questionnaire data capture April-May 2020. Wellcome Open Res. 2020;5:127.

47. Kann L, Warren CW, Harris WA, Collins JL, Williams BI, Ross JG, et al. Youth Risk Behavior Surveillance-United States, 1995. J Sch Health. 1996;66(10):365–77.

48. APA. Diagnostic and statistical manual of mental disorders. 5th ed. Washington, DC: American Psychiatric Association; 2013.

49. Madge N, Hewitt A, Hawton K, Wilde EJ De, Corcoran P, Fekete S, et al. Deliberate self-harm within an international community sample of young people: Comparative findings from the Child & Adolescent Self-harm in Europe (CASE) Study. J Child Psychol Psychiatry Allied Discip. 2008;49(6):667–77.

50. Angold A, Costello EJ, Messer SC, Pickles A, Winder F, Silver D. The development of a short questionnaire for use in epidemiological studies of depression in children and adolescents. Int J Methods Psychiatr Res. 1995;5(5):237–49.

51. Flora DB. Your Coefficient Alpha Is Probably Wrong, but Which Coefficient Omega Is Right? A Tutorial on Using R to Obtain Better Reliability Estimates: https://doi.org/101177/2515245920951747 [Internet]. 2020 Nov 6 [cited 2021 Aug 24];3(4):484–501. Available from: https://journals.sagepub.com/doi/full/10.1177/2515245920951747

52. Rosseel Y. lavaan: An R package for structural equation modeling. J Stat Softw. 2012;48(2).

53. Spitzer RL, Kroenke K, Williams JBW, Löwe B. A brief measure for assessing generalized anxiety disorder: The GAD-7. Arch Intern Med. 2006;166(10):1092–7.

54. Tennant R, Hiller L, Fishwick R, Platt S, Joseph S, Weich S, et al. The Warwick-Edinburgh mental well-being scale (WEMWBS): Development and UK validation. Health Qual Life Outcomes. 2007;5(1):63.

55. StataCorp. Stata Statistical Software: Release 16. College Station, TX: StataCorp LLC; 2019.

56. Royston P, White IR. Multiple Imputation by Chained Equations (MICE): Implementation in Stata. J Stat Softw. 2011;45(4):1–20.

57. Tilling K, Williamson EJ, Spratt M, Sterne JAC, Carpenter JR. Appropriate inclusion of interactions was needed to avoid bias in multiple imputation. J Clin Epidemiol. 2016 Dec 1;80:107–15.

58. Mansfield KE, Mathur R, Tazare J, Henderson AD, Mulick AR, Carreira H, et al. Indirect acute effects of the COVID-19 pandemic on physical and mental health in the UK: a population-based study. Lancet Digit Heal. 2021;3(4):e217–30.

59. Parker LL, Harriger JA. Eating disorders and disordered eating behaviors in the LGBT population: a review of the literature. J Eat Disord 2020 81 [Internet]. 2020 Oct 16 [cited 2021 Nov 1];8(1):1–20. Available from: https://jeatdisord.biomedcentral.com/articles/10.1186/s40337-020-00327-y

60. Marshall E, Claes L, Bouman WP, Witcomb GL, Arcelus J. Non-suicidal self-injury and suicidality in trans people: A systematic review of the literature. https://doi.org/103109/0954026120151073143 [Internet]. 2015 Jan 2 [cited 2021 Nov 1];28(1):58–69. Available from: https://www.tandfonline.com/doi/abs/10.3109/09540261.2015.1073143

61. Kidd JD, Jackman KB, Barucco R, Dworkin JD, Dolezal C, Navalta T V., et al. Understanding the Impact of the COVID-19 Pandemic on the Mental Health of Transgender and Gender Nonbinary Individuals Engaged in a Longitudinal Cohort Study. J Homosex [Internet]. 2021 [cited 2021 Nov 11];68(4):592–611. Available from: https://pubmed.ncbi.nlm.nih.gov/33502286/

