## Supplementary for "Disordered eating and self-harm as risk factors for poorer mental health during the COVID-19 pandemic: A UK-based birth cohort study"

Supplementary Table 1. Comparison of responders and non-responders to the lifestyle change questions on the COVID1 questionnaire

| **Variable** | **Description** | **n** | **Data on lifestyle changes** | **No data on lifestyle changes** | **$\chi^2$** | **p** |
|---|---|---|---|---|---|---|
| Child gender | Male | 2053 | 766 (28.83%) | 1287 (48.11%) | 209.32 | <.001 |
| | Female | 3279 | 1891 (71.17%) | 1388 (51.89%) | | |
| Child ethnicity | White | 4865 | 2455 (96.58%) | 2410 (95.45%) | 4.24 | 0.039 |
| | Not white | 202 | 87 (3.42%) | 115 (4.55%) | | |
| NEET at 23 | In education, employment, or training | 2967 | 1886 (94.96) | 1081 (92.79%) | 6.32 | 0.012 |
| | NEET | 184 | 100 (5.04%) | 84 (7.21%) | | |
| Maternal education | O level/GCSE and below | 2,723 | 1289 (50.08%) | 1434 (55.88%) | 17.39 | <.001 |
| | A level and above (degree) | 2,417 | 1285 (49.92%) | 1132 (44.12%) | | |
| Parity | First born | 2,466 | 1267 (49.11%) | 1199 (46.69%) | 4.9 | 0.086 |
| | Second born | 1,794 | 895 (34.69%) | 899 (35.01%) | | |
| | Third or more born | 888 | 418 (16.20%) | 470 (18.30%) | | |
| Home ownership | Don't own home | 865 | 377 (14.58% | 488 (18.91%) | 17.37 | <.001 |
| | Own home | 4,302 | 2209 (85.42%) | 1093 (81.09%) | | |

Comparison amongst those sent the COVID1 questionnaire (n=5332). NEET = Not in education, employment, or training

Supplementary Table 2. Derivation of disordered eating and self-harm variables at age 24

| Questions | Answers | Coding | Final variable |
|---|---|---|---|
| **Fasting** | | | |
| During the **<u>past year</u>**, how often did you fast (not eat for at least a day) to lose weight or avoid gaining weight? | 0. Never<br>1. Less than once a month<br>2. 1-3 times a month<br>3. Once a week<br>4. More than once a week | Any fasting<br>0 = "no"<br>1-4 = "yes"<br>DSM fasting<br>0-2 = "no"<br>3-4 = "yes" | **Any fasting** |
| **Purging** | | | |
| During the **<u>past year</u>**, how often did you make yourself throw up to lose weight or avoid gaining weight? | 0. Never<br>1. Less than once a month<br>2. 1-3 times a month<br>3. Once a week<br>4. More than once a week | Any self-induced vomiting<br>0 = "no"<br>1-4 = "yes"<br>DSM self-induced vomiting<br>0-2 = "no"<br>3-4 = "yes" | **Any purging**<br>Any self-induced vomiting<br>OR<br>Any laxative use<br>OR<br>Any other medication use |
| During the **<u>past year</u>**, how often did you take laxatives to lose weight or avoid gaining weight? | 0. Never<br>1. Less than once a month<br>2. 1-3 times a month<br>3. Once a week<br>4. More than once a week | Any laxative use<br>0 = "no"<br>1-4 = "yes"<br>DSM laxative use<br>0-2 = "no"<br>3-4 = "yes" | |
| During the **<u>past year</u>**, how often did you take other tablets/pills/any other medications or substances to lose weight or avoid gaining weight? | 0. Never<br>1. Less than once a month<br>2. 1-3 times a month<br>3. Once a week<br>4. More than once a week | Any other medication use<br>0 = "no"<br>1-4 = "yes"<br>DSM other medication use<br>0-2 = "no"<br>3-4 = "yes" | |
| **Excessive exercise** | | | |
| During the **<u>past year</u>**, how often did you exercise to **<u>lose weight</u>** or **<u>avoid gaining weight</u>**? | 0. Never<br>1. Less than once a month<br>2. 1-3 times a month<br>3. 1-4 times a week<br>4. 5 or more times a week | Any exercise to lose weight<br>0 = "no"<br>1-4 = "yes"<br>DSM exercise to lose weight<br>0-2 = "no"<br>3-4 = "yes" | **Any excessive exercise**<br>Any exercise to lose weight<br>AND<br><br>Exercise interfered with life<br>OR<br>Exercise when sick/injured |
| Was it difficult for you to do your work or daily chores/routine because of the amount of time that you were exercising to lose weight or avoid gaining weight? | 0. No<br>1. Yes, sometimes<br>2. Yes, frequently | Exercise interfered with life<br>0-1 = "no"<br>2 = "yes" | |
| Did you exercise to lose weight or avoid gaining weight even when you were sick or injured? | 0. No<br>1. Yes, sometimes<br>2. Yes, frequently | Exercise when sick/injured<br>0-1 = "no"<br>2 = "yes" | |
| **Binge eating** | | | |
| Sometimes people will go on an 'eating binge', where they eat an amount of food that most people, like their friends | 0. Never<br>1. Less than once a month<br>2. 1-3 times a month<br>3. Once a week | Any bingeing<br>0 = "no"<br>1-4 = "yes"<br><br>DSM bingeing | **Any binge eating**<br>Any bingeing<br>AND<br>Loss of control |

| | | | |
|---|---|---|---|
| or family, would consider to be very large in a short period of time. During the **past year**, how often did you go on an eating binge? | 4. More than once a week | 0-2 = "no"<br>3-4 = "yes" | |
| Do you ever feel like your eating is out of control, like you couldn't stop eating even if you wanted to? | 0. No<br>1. Yes, sometimes<br>2. Yes, usually | Loss of control<br>0 = "no"<br>1-2 = "yes" | |
| **Any disordered eating** | | | |
| | | | **Any disordered eating**<br>Any fasting OR<br>Any purging OR<br>Any binge-eating OR<br>Any excessive exercise |
| **DSM-5 frequency disordered eating** | | | |
| | | **DSM frequency purging**<br>DSM self-induced vomiting OR<br>DSM laxative use OR<br>DSM Other medication use<br><br>**DSM frequency binge-eating**<br>DSM bingeing AND<br>Loss of control<br><br>**DSM frequency excessive exercise**<br>DSM exercise to lose weight AND<br>Exercise interfered with life | **DSM-5 frequency disordered eating**<br>DSM frequency fasting OR<br>DSM frequency purging OR<br>DSM frequency binge-eating<br>OR<br>DSM frequency excessive exercise |
| **Self-harm for any reason** | | | |
| Have you **ever** hurt yourself on purpose in any way (e.g. by taking an overdose of pills or by cutting yourself)? | 0. No<br>1. Yes | *Self-harm ever*<br>0 = "no"<br>1 = "yes" | **Self-harm in the last year**<br>*Self-harm past year*<br>(= 0 if self-harm ever = 0) |
| **If yes,** how many times have you done this in the last year? | 0. None<br>1. Once<br>2. 2-5 times<br>3. 6-10 times<br>4. More than 10 times | *Self-harm past year*<br>0 = "no"<br>1-4 = "yes" | |
| **Non-suicidal self-injury** | | | |
| Have you **ever** hurt yourself on purpose (e.g. by taking an overdose of pills or by cutting yourself), without intending to kill yourself? | 0. No<br>1. Yes | *NSSI ever*<br>0 = "no"<br>1 = "yes" | **NSSI in the last year**<br>*NSSI past year*<br>(= 0 if self-harm ever OR NSSI ever = 0) |

| | | | |
|---|---|---|---|
| If yes, when was the last time you hurt yourself on purpose, without intending to kill yourself? | 1. In the last week<br>2. More than a week ago but in the last year<br>3. More than a year ago | *NSSI past year*<br>3 = "no"<br>1-2 = "yes" | |
| **Suicide attempt** | | | |
| On any of the occasions you have hurt yourself on purpose, have you **ever** seriously wanted to kill yourself? | 0. No<br>1. Yes | *Attempt to kill ever*<br>0 = "no"<br>1 = "yes" | Suicide attempt ever<br>*Attempt to kill ever*<br>OR<br>*Reason: to die* |
| **In your lifetime,** do any of the following reasons help to explain why you hurt yourself? | a) I wanted to show how desperate I was feeling Y/N<br>b) I wanted to die<br>c) I wanted to punish myself<br>d) I wanted to frighten someone<br>e) I wanted to get relief from a terrible state of mind<br>f) Some other reason | *Reason: to die*<br>b 0 = "no"<br>b 1 = "yes" | |
| **If yes**, when was the last time you hurt yourself on purpose and you seriously wanted to kill yourself? | 1. In the last week<br>2. More than a week ago but in the last year<br>3. More than a year ago | *Suicide attempt past year*<br>3 = "no"<br>1-2 = "yes" | **Suicide attempt in the last year**<br>*Suicide attempt past year*<br>(= 0 if suicide attempt ever = 0) |

Supplementary Table 3. Question wording and variable coding for pre-pandemic measure of socioeconomic disadvantage (young person not in education, employment or training)

| **Question wording** | | **Possible responses** | **Final variable coding** |
|---|---|---|---|
| Are you currently | | | |
| | In full-time paid work (30 hours or more a week) | Yes (1)<br>No (0) | If all responses are No (0), then NEET variable is 1 (yes)<br><br>If any response is Yes (1), then NEET variable is 0 (No) |
| | In part-time paid work (less than 30 hours a week) | Yes (1)<br>No (0) | |
| | In irregular or occasional work | Yes (1)<br>No (0) | |
| | Doing a modern apprenticeship or other government supported training/work-experience scheme | Yes (1)<br>No (0) | |
| | In full-time education | Yes (1)<br>No (0) | |
| | Self-employed | Yes (1)<br>No (0) | |

Supplementary Table 4. Question wording and variable coding for pandemic-related experiences

| **Variable** | **Questionnaire Wording** | **Response Options** | **Final variable coding** |
|---|---|---|---|
| Living alone | Do you live with anybody? | No I live on my own<br>Yes, I live with at least 1 person | If "No I live on my own" then *living alone* = Yes (1) |
| Keyworker | Are you a keyworker, or has your work been classified as critical to the COVID-19 response? | Yes<br>No<br>Don't know | If "Yes" then *keyworker* = Yes (1), If "No" then *keyworker* = No (0)<br>If "Don't know" then *keyworker* = missing |
| Financial problems during the pandemic | Overall, how do you feel your current financial situation compares to how it was before the COVID-19 pandemic? | I'm much worse off<br>I'm a little worse off<br>I'm about the same<br>I'm a little better off<br>I'm much better off | If "I'm much worse off" or "I'm a little worse off" then *financial problems* = Yes (1)<br>Any other response then *financial problems* = No (0) |
| Furloughed during the pandemic | Which of these would you say best describes your current situation now? | Employed and working the same number of hours (as pre-lockdown)<br>Employed and working reduced number of hours<br>Employed and working more hours than before<br>Employed but on paid leave (including furlough)<br>Employed and on unpaid leave<br>Apprenticeship<br>In unpaid/voluntary work<br>Self-employed and currently working<br>Self-employed but not currently working<br>Unemployed<br>Permanently sick or disabled<br>Looking after home or family<br>In education at school/college/university | If "Employed but on paid leave (including furlough)" then *furlough* = Yes (1)<br>Any other response then *furlough* = No (0) |

Supplementary Table 5. Amount of missing data on exposures, outcomes and confounders

| **Variable** | **n present** | **n missing** | **% missing** |
|---|---|---|---|
| *Primary exposures* | | | |
| Any disordered eating | 2096 | 561 | 21.11% |
| Any self-harm | 2115 | 542 | 20.40% |
| Comorbid disordered eating and self-harm | 2089 | 568 | 21.38% |
| *Secondary exposures* | | | |
| Fasting | 2115 | 542 | 20.40% |
| Purging | 2112 | 545 | 20.51% |
| Binge-eating | 2116 | 541 | 20.36% |
| Excessive exercise | 2105 | 552 | 20.78% |
| DSM-5 frequency disordered eating | 2093 | 564 | 21.23% |
| Self-harm without suicidal intent | 2115 | 542 | 20.40% |
| Self-harm with suicidal intent | 2081 | 576 | 21.68% |
| *Outcomes* | | | |
| Depressive symptoms | 1914 | 743 | 27.96% |
| Anxiety symptoms | 1916 | 741 | 27.89% |
| Mental wellbeing | 1922 | 735 | 27.66% |
| *Confounders* | | | |
| Gender | 2657 | 0 | 0.00% |
| COVID1 questionnaire completion date | 2657 | 0 | 0.00% |
| Pre-pandemic NEET | 1986 | 671 | 25.25% |
| Pre-pandemic depressive symptoms | 1998 | 659 | 24.80% |
| Pre-pandemic anxiety symptoms | 1718 | 939 | 35.34% |
| Pre-pandemic mental wellbeing | 2014 | 643 | 24.20% |

NEET = Not in employment, education or training

Supplementary Table 6. Comparison of complete case and imputed analysis for main effects of disordered eating and self-harm on unstandardised pandemic mental health outcomes

| | | | Complete case sample 1 | | | Complete case sample 2 | | | Imputed data | | |
|---|---|---|---|---|---|---|---|---|---|---|---|
| | | | n | B (95% CI) | p | n | B (95% CI) | p | n | B (95% CI) | p |
| Outcome: depressive symptoms | DE | Model A | 1341 | 3.26 (2.62, 3.90) | <.001 | 1585 | 3.09 (2.51, 3.68) | <.001 | 2657 | 2.98 (2.44, 3.53) | <.001 |
| | | Model B | | | | 1360 | 2.95 (2.31, 3.59) | <.001 | 2657 | 2.72 (2.17, 3.27) | <.001 |
| | | Model C | | | | 1341 | 1.48 (0.88, 2.09) | <.001 | 2657 | 1.37 (0.84, 1.90) | <.001 |
| | SH | Model A | 1350 | 5.37 (4.33, 6.41) | <.001 | 1594 | 5.29 (4.36, 6.22) | <.001 | 2657 | 5.19 (4.31, 6.08) | <.001 |
| | | Model B | | | | 1369 | 5.20 (4.18, 6.21) | <.001 | 2657 | 4.95 (4.07, 5.83) | <.001 |
| | | Model C | | | | 1350 | 2.34 (1.36, 3.33) | <.001 | 2657 | 2.13 (1.24, 3.01) | <.001 |
| | DE+SH | Model A | 1337 | 5.89 (4.59, 7.18) | <.001 | 1580 | 5.91 (4.72, 7.10) | <.001 | 2657 | 6.15 (5.02, 7.29) | <.001 |
| | | Model B | | | | 1356 | 5.61 (4.35, 6.87) | <.001 | 2657 | 5.87 (4.74, 7.00) | <.001 |
| | | Model C | | | | 1337 | 2.22 (1.01, 3.44) | <.001 | 2657 | 2.52 (1.38, 3.66) | <.001 |
| Outcome: Anxiety symptoms | DE | Model A | 1094 | 2.59 (1.97, 3.22) | <.001 | 1586 | 2.50 (1.97, 3.04) | <.001 | 2657 | 2.43 (1.92, 2.95) | <.001 |
| | | Model B | | | | 1359 | 2.32 (1.74, 2.90) | <.001 | 2657 | 2.11 (1.59, 2.63) | <.001 |
| | | Model C | | | | 1094 | 1.26 (0.66, 1.86) | <.001 | 2657 | 1.24 (0.74, 1.74) | <.001 |
| | SH | Model A | 1100 | 4.02 (3.03, 5.01) | <.001 | 1595 | 4.46 (3.62, 5.31) | <.001 | 2657 | 4.55 (3.74, 5.36) | <.001 |
| | | Model B | | | | 1368 | 4.05 (3.13, 4.96) | <.001 | 2657 | 4.26 (3.46, 5.05) | <.001 |
| | | Model C | | | | 1100 | 2.13 (1.19, 3.06) | <.001 | 2657 | 2.69 (1.87, 3.50) | <.001 |
| | DE+SH | Model A | 1090 | 4.52 (3.28, 5.75) | <.001 | 1581 | 5.09 (4.02, 6.17) | <.001 | 2657 | 5.38 (4.30, 6.47) | <.001 |
| | | Model B | | | | 1355 | 4.52 (3.39, 5.64) | <.001 | 2657 | 5.05 (3.98, 6.12) | <.001 |
| | | Model C | | | | 1090 | 2.27 (1.11, 3.43) | <.001 | 2657 | 3.08 (2.01, 4.15) | <.001 |
| Outcome: mental wellbeing | DE | Model A | 1343 | -3.71 (-4.67, -2.75) | <.001 | 1592 | -3.66 (-4.55, -2.77) | <.001 | 2657 | -3.49 (-4.30, -2.67) | <.001 |
| | | Model B | | | | 1364 | -3.72 (-4.69, -2.74) | <.001 | 2657 | -3.35 (-4.18, -2.51) | <.001 |
| | | Model C | | | | 1343 | -1.75 (-2.65, -0.85) | <.001 | 2657 | -1.82 (-2.59, -1.06) | <.001 |
| | SH | Model A | 1351 | -6.31 (-7.88, -4.73) | <.001 | 1601 | -6.10 (-7.54, -4.67) | <.001 | 2657 | -5.78 (-7.06, -4.50) | <.001 |
| | | Model B | | | | 1373 | -6.14 (-7.70, -4.58) | <.001 | 2657 | -5.62 (-6.90, -4.33) | <.001 |
| | | Model C | | | | 1351 | -2.42 (-3.89, -0.94) | 0.001 | 2657 | -2.18 (-3.43, -0.93) | <.001 |
| | DE+SH | Model A | 1339 | -8.09 (-10.02, -6.15) | <.001 | 1587 | -8.10 (-9.91, -6.30) | <.001 | 2657 | -7.81 (-9.43, -6.19) | <.001 |
| | | Model B | | | | 1360 | -7.79 (-9.70, -5.87) | <.001 | 2657 | -7.60 (-9.21, -6.00) | <.001 |
| | | Model C | | | | 1339 | -3.45 (-5.26, -1.65) | <.001 | 2657 | -3.64 (-5.19, -2.09) | <.001 |

Complete case sample 1 = Individuals with complete data on exposure, outcome and all confounders in the final model (n varies by model); Complete case sample 2 = all available observed data for each model (n varies by model); Imputed data = individuals with complete data on lifestyle changes with imputed data for exposures, outcomes and confounders (n=2657) (see manuscript Missing Data section).
DE = disordered eating; SH = Self-harm; DE+SH = comorbid disordered eating and self-harm.
Model A = unadjusted; Model B = adjusted for sex, COVID1 questionnaire completion date, pre-pandemic socioeconomic status; Model C = adjusted for sex, COVID1 questionnaire completion date, pre-pandemic socioeconomic status and pre-pandemic mental health and wellbeing symptoms.

Supplementary Table 7. Associations between secondary disordered eating and self-harm exposures and mental health outcomes during the pandemic

| | Unadjusted Model A | | | Adjusted Model B | | | Fully adjusted Model C | | |
|---|---|---|---|---|---|---|---|---|---|
| **Exposure** | **n** | **B (95% CI)** | **p** | **n** | **B (95% CI)** | **p** | **n** | **B (95% CI)** | **p** |
| ***Outcome: Depressive symptoms*** | | | | | | | | | |
| Fasting | 1595 | 3.73 (2.83, 4.63) | <.001 | 1370 | 3.25 (2.27, 4.23) | <.001 | 1351 | 1.60 (0.70, 2.50) | <.001 |
| Purging | 1593 | 3.56 (2.60, 4.51) | <.001 | 1367 | 3.39 (2.35, 4.44) | <.001 | 1348 | 1.84 (0.89, 2.79) | <.001 |
| Binge-eating | 1596 | 2.68 (2.00, 3.36) | <.001 | 1370 | 2.57 (1.84, 3.30) | <.001 | 1351 | 1.25 (0.58, 1.92) | <.001 |
| Excessive exercise | 1592 | 2.17 (0.66, 3.69) | 0.005 | 1368 | 1.95 (0.26, 3.64) | 0.024 | 1349 | 0.75 (-0.75, 2.25) | 0.324 |
| DSM-5 frequency disordered eating | 1583 | 3.75 (2.81, 4.69) | <.001 | 1360 | 3.58 (2.55, 4.61) | <.001 | 1341 | 1.93 (0.99, 2.87) | <.001 |
| Self-harm without suicidal intent | 1593 | 4.57 (3.48, 5.67) | <.001 | 1368 | 4.42 (3.24, 5.60) | <.001 | 1349 | 1.46 (0.35, 2.57) | 0.01 |
| Self-harm with suicidal intent | 1569 | 8.00 (5.95, 10.05) | <.001 | 1349 | 9.22 (7.05, 11.39) | <.001 | 1330 | 4.88 (2.88, 6.88) | <.001 |
| ***Outcome: Anxiety symptoms*** | | | | | | | | | |
| Fasting | 1595 | 3.13 (2.31, 3.96) | <.001 | 1368 | 2.56 (1.67, 3.44) | <.001 | 1101 | 0.99 (0.08, 1.89) | 0.032 |
| Purging | 1593 | 2.81 (1.93, 3.69) | <.001 | 1365 | 2.29 (1.34, 3.24) | <.001 | 1099 | 1.35 (0.42, 2.28) | 0.005 |
| Binge-eating | 1596 | 2.08 (1.47, 2.70) | <.001 | 1368 | 1.97 (1.32, 2.63) | <.001 | 1100 | 1.19 (0.53, 1.86) | <.001 |
| Excessive exercise | 1593 | 1.81 (0.44, 3.19) | 0.01 | 1367 | 1.65 (0.15, 3.14) | 0.031 | 1099 | 1.79 (0.21, 3.36) | 0.026 |
| DSM-5 frequency disordered eating | 1584 | 2.85 (1.99, 3.70) | <.001 | 1359 | 2.79 (1.87, 3.72) | <.001 | 1094 | 1.67 (0.71, 2.62) | 0.001 |
| Self-harm without suicidal intent | 1594 | 3.47 (2.48, 4.46) | <.001 | 1367 | 3.21 (2.17, 4.26) | <.001 | 1100 | 1.70 (0.68, 2.73) | 0.001 |
| Self-harm with suicidal intent | 1570 | 5.63 (3.73, 7.54) | <.001 | 1348 | 6.56 (4.57, 8.54) | <.001 | 1082 | 3.77 (1.73, 5.81) | <.001 |
| ***Outcome: Mental wellbeing*** | | | | | | | | | |
| Fasting | 1601 | -4.76 (-6.11, -3.41) | <.001 | 1373 | -4.22 (-5.69, -2.74) | <.001 | 1351 | -1.76 (-3.10, -0.41) | 0.011 |
| Purging | 1599 | -3.81 (-5.27, -2.35) | <.001 | 1370 | -3.74 (-5.33, -2.15) | <.001 | 1349 | -1.49 (-2.93, -0.06) | 0.041 |
| Binge-eating | 1602 | -3.36 (-4.39, -2.34) | <.001 | 1373 | -3.30 (-4.41, -2.20) | <.001 | 1351 | -1.63 (-2.63, -0.63) | 0.001 |
| Excessive exercise | 1599 | -1.80 (-4.07, 0.47) | 0.12 | 1372 | -1.48 (-4.00, 1.04) | 0.25 | 1350 | -0.19 (-2.49, 2.12) | 0.875 |
| DSM-5 frequency disordered eating | 1590 | -4.14 (-5.57, -2.71) | <.001 | 1364 | -3.97 (-5.54, -2.40) | <.001 | 1343 | -1.82 (-3.25, -0.39) | 0.013 |
| Self-harm without suicidal intent | 1600 | -5.10 (-6.77, -3.43) | <.001 | 1372 | -5.16 (-6.95, -3.37) | <.001 | 1350 | -1.28 (-2.93, 0.37) | 0.127 |
| Self-harm with suicidal intent | 1576 | -10.28 (-13.45, -7.10) | <.001 | 1353 | -12.20 (-15.57, -8.84) | <.001 | 1331 | -6.46 (-9.58, -3.33) | <.001 |

Analyses conducted using all available observed data for each model. Model A = unadjusted; Model B = adjusted for sex, COVID1 questionnaire completion date, pre-pandemic socioeconomic status; Model C = adjusted for sex, COVID1 questionnaire completion date, pre-pandemic socioeconomic status and pre-pandemic mental health and wellbeing symptoms.

Supplementary Table 8. Associations between disordered eating frequency exposures and mental health outcomes during the pandemic

| | **Unadjusted Model A** | | | **Adjusted Model B** | | | **Fully adjusted Model C** | | |
|---|---|---|---|---|---|---|---|---|---|
| **Exposure** | **n** | **B (95% CI)** | **p** | **n** | **B (95% CI)** | **p** | **n** | **B (95% CI)** | **p** |
| ***Outcome: Depressive symptoms*** | | | | | | | | | |
| Disordered eating less than once a week | 1585 | 2.54 (1.87, 3.20) | <.001 | 1360 | 2.43 (1.71, 3.15) | <.001 | 1341 | 1.17 (0.50, 1.83) | 0.001 |
| Disordered eating once a week or more | | 4.36 (3.42, 5.29) | <.001 | | 4.25 (3.21, 5.28) | <.001 | | 2.31 (1.35, 3.27) | <.001 |
| ***Outcome: Anxiety symptoms*** | | | | | | | | | |
| Disordered eating less than once a week | 1586 | 2.13 (1.52, 2.74) | <.001 | 1359 | 1.91 (1.27, 2.56) | <.001 | 1094 | 0.99 (0.33, 1.66) | 0.003 |
| Disordered eating once a week or more | | 3.35 (2.49, 4.21) | <.001 | | 3.32 (2.38, 4.25) | <.001 | | 1.97 (1.00, 2.95) | <.001 |
| ***Outcome: Mental wellbeing*** | | | | | | | | | |
| Disordered eating less than once a week | 1592 | -3.13 (-4.15, -2.12) | <.001 | 1364 | -3.25 (-4.35, -2.16) | <.001 | 1343 | -1.54 (-2.54, -0.54) | 0.002 |
| Disordered eating once a week or more | | -4.88 (-6.31, -3.45) | <.001 | | -4.87 (-6.45, -3.29) | <.001 | | -2.31 (-3.77, -0.85) | 0.002 |

Analyses conducted using all available observed data for each model. Reference category is no disordered eating (0).

Model A = unadjusted; Model B = adjusted for sex, COVID1 questionnaire completion date, pre-pandemic socioeconomic status; Model C = adjusted for sex, COVID1 questionnaire completion date, pre-pandemic socioeconomic status and pre-pandemic mental health and wellbeing symptoms.

Supplementary Table 9. Lifestyle changes during lockdown (n=2657)

| **Since lockdown the participant has changed the amount:** | **Decrease** | | **Stayed the same** | | **Increase** | |
|---|---|---|---|---|---|---|
| | n | % | n | % | n | % |
| they *sleep* | 628 | 23.64 | 1,152 | 43.36 | 877 | 33.01 |
| of physical activity/*exercise* | 1,151 | 43.32 | 697 | 26.23 | 809 | 30.45 |
| of *alcohol* drunk | 397 | 14.94 | 1,362 | 51.26 | 898 | 33.80 |
| visiting *green space* | 1,207 | 45.43 | 692 | 26.04 | 758 | 28.53 |
| they *eat* | 241 | 9.07 | 1,319 | 49.64 | 1,097 | 41.29 |
| of time spent *talking* to family/friends outside their home | 982 | 36.96 | 524 | 19.72 | 1,151 | 43.32 |
| of time spent doing *hobbies*/things they enjoy | 478 | 17.99 | 891 | 33.53 | 1,288 | 48.48 |
| of practising *relaxation*/mindfulness/meditation | 136 | 5.12 | 2,021 | 76.06 | 500 | 18.82 |

Supplementary Table 10. Associations between disordered eating/self-harm exposures (age 25 years, YPD questionnaire) and lifestyle change moderators (COVID1 questionnaire)

| **Exposure** | **Lifestyle change moderator** | **Decreased (ref = stayed same)** | | **Increased (ref = stayed same)** | | |
|---|---|---|---|---|---|---|
| | | **RRR** | **95% CI** | **RRR** | **95% CI** | **p** |
| Disordered eating | Sleep | 2.055 | [1.630, 2.589] | 1.568 | [1.267, 1.939] | < 0.0001 |
| Disordered eating | Exercise | 1.214 | [0.974, 1.511] | 1.039 | [0.818, 1.320] | 0.1592 |
| Disordered eating | Alcohol | 0.971 | [0.744, 1.267] | 1.292 | [1.062, 1.573] | 0.0217 |
| Disordered eating | Green space | 1.333 | [1.062, 1.673] | 1.098 | [0.850, 1.419] | 0.0294 |
| Disordered eating | Eating | 1.732 | [1.251, 2.398] | 1.881 | [1.547, 2.286] | < 0.0001 |
| Disordered eating | Talking to friends/family | 1.170 | [0.903, 1.516] | 1.117 | [0.871, 1.433] | 0.4783 |
| Disordered eating | Hobbies | 1.080 | [0.828, 1.408] | 1.111 | [0.906, 1.363] | 0.6001 |
| Disordered eating | Relaxation | 1.335 | [0.900, 1.979] | 1.123 | [0.887, 1.422] | 0.2642 |
| Self-harm | Sleep | 1.658 | [1.165, 2.361] | 1.056 | [0.741, 1.505] | 0.0109 |
| Self-harm | Exercise | 1.138 | [0.805, 1.610] | 0.711 | [0.473, 1.068] | 0.0400 |
| Self-harm | Alcohol | 0.884 | [0.569, 1.372] | 0.966 | [0.703, 1.328] | 0.8574 |
| Self-harm | Green space | 1.100 | [0.777, 1.556] | 0.741 | [0.487, 1.129] | 0.1109 |
| Self-harm | Eating | 1.719 | [1.059, 2.791] | 1.230 | [0.895, 1.689] | 0.0759 |
| Self-harm | Talking to friends/family | 1.362 | [0.874, 2.122] | 1.325 | [0.861, 2.040] | 0.3536 |
| Self-harm | Hobbies | 1.189 | [0.793, 1.783] | 0.912 | [0.660, 1.261] | 0.3984 |
| Self-harm | Relaxation | 2.489 | [1.469, 4.219] | 1.098 | [0.753, 1.602] | 0.0026 |

Results using imputed data (n=2657).

Supplementary Table 11. Unadjusted associations between lifestyle change moderators (COVID1 questionnaire) and pandemic mental health and wellbeing outcomes (COVID2 questionnaire)

| **Lifestyle change** | **Mental health outcome** | **Decreased (ref = stayed same)** | | **Increased (ref = stayed same)** | | |
|---|---|---|---|---|---|---|
| | | **Diff (SE(Diff))** | **95% CI** | **Diff (SE(Diff))** | **95% CI** | **p** |
| Sleep | Depressive symptoms | 0.564 (0.052) | (0.462, 0.666) | 0.224 (0.047) | (0.132, 0.317) | < 0.0001 |
| Exercise | Depressive symptoms | 0.234 (0.051) | (0.134, 0.333) | 0.015 (0.056) | (-0.094, 0.124) | < 0.0001 |
| Alcohol | Depressive symptoms | -0.007 (0.060) | (-0.126, 0.111) | 0.101 (0.047) | (0.009, 0.194) | 0.0655 |
| Green space | Depressive symptoms | 0.235 (0.050) | (0.136, 0.333) | 0.065 (0.056) | (-0.044, 0.174) | < 0.0001 |
| Eating | Depressive symptoms | 0.431 (0.075) | (0.283, 0.579) | 0.231 (0.043) | (0.146, 0.317) | < 0.0001 |
| Talking to friends/family | Depressive symptoms | 0.114 (0.058) | (0.001, 0.226) | 0.064 (0.057) | (-0.047, 0.175) | 0.1426 |
| Hobbies | Depressive symptoms | 0.165 (0.061) | (0.045, 0.285) | -0.030 (0.047) | (-0.123, 0.063) | 0.0033 |
| Relaxation | Depressive symptoms | 0.500 (0.098) | (0.307, 0.693) | 0.134 (0.055) | (0.026, 0.241) | < 0.0001 |
| Sleep | Anxiety symptoms | 0.662 (0.052) | (0.560, 0.764) | 0.252 (0.046) | (0.161, 0.342) | < 0.0001 |
| Exercise | Anxiety symptoms | 0.154 (0.051) | (0.053, 0.255) | 0.053 (0.056) | (-0.056, 0.162) | 0.0074 |
| Alcohol | Anxiety symptoms | 0.011 (0.060) | (-0.107, 0.129) | 0.045 (0.047) | (-0.046, 0.137) | 0.6121 |
| Green space | Anxiety symptoms | 0.199 (0.051) | (0.099, 0.300) | 0.080 (0.056) | (-0.029, 0.190) | 0.0003 |
| Eating | Anxiety symptoms | 0.513 (0.074) | (0.368, 0.659) | 0.232 (0.044) | (0.146, 0.318) | < 0.0001 |
| Talking to friends/family | Anxiety symptoms | 0.128 (0.058) | (0.014, 0.242) | 0.118 (0.057) | (0.006, 0.229) | 0.0641 |
| Hobbies | Anxiety symptoms | 0.133 (0.061) | (0.013, 0.252) | -0.025 (0.046) | (-0.116, 0.066) | 0.0216 |
| Relaxation | Anxiety symptoms | 0.548 (0.095) | (0.362, 0.734) | 0.236 (0.054) | (0.130, 0.341) | < 0.0001 |
| Sleep | Mental wellbeing | -0.540 (0.053) | (-0.644, -0.437) | -0.095 (0.048) | (-0.189, -0.002) | < 0.0001 |
| Exercise | Mental wellbeing | -0.192 (0.051) | (-0.292, -0.093) | 0.061 (0.056) | (-0.048, 0.170) | < 0.0001 |
| Alcohol | Mental wellbeing | -0.004 (0.061) | (-0.123, 0.116) | -0.086 (0.047) | (-0.177, 0.005) | 0.1568 |
| Green space | Mental wellbeing | -0.208 (0.052) | (-0.310, -0.107) | -0.028 (0.057) | (-0.140, 0.083) | < 0.0001 |
| Eating | Mental wellbeing | -0.264 (0.074) | (-0.410, -0.118) | -0.222 (0.045) | (-0.309, -0.134) | < 0.0001 |
| Talking to friends/family | Mental wellbeing | -0.049 (0.058) | (-0.163, 0.065) | 0.043 (0.057) | (-0.069, 0.156) | 0.1441 |
| Hobbies | Mental wellbeing | -0.083 (0.061) | (-0.203, 0.036) | 0.191 (0.048) | (0.097, 0.285) | < 0.0001 |
| Relaxation | Mental wellbeing | -0.469 (0.097) | (-0.658, -0.279) | 0.099 (0.055) | (-0.009, 0.206) | < 0.0001 |

Results using imputed data (n=2657).

Supplementary Figure 1. Participant attrition

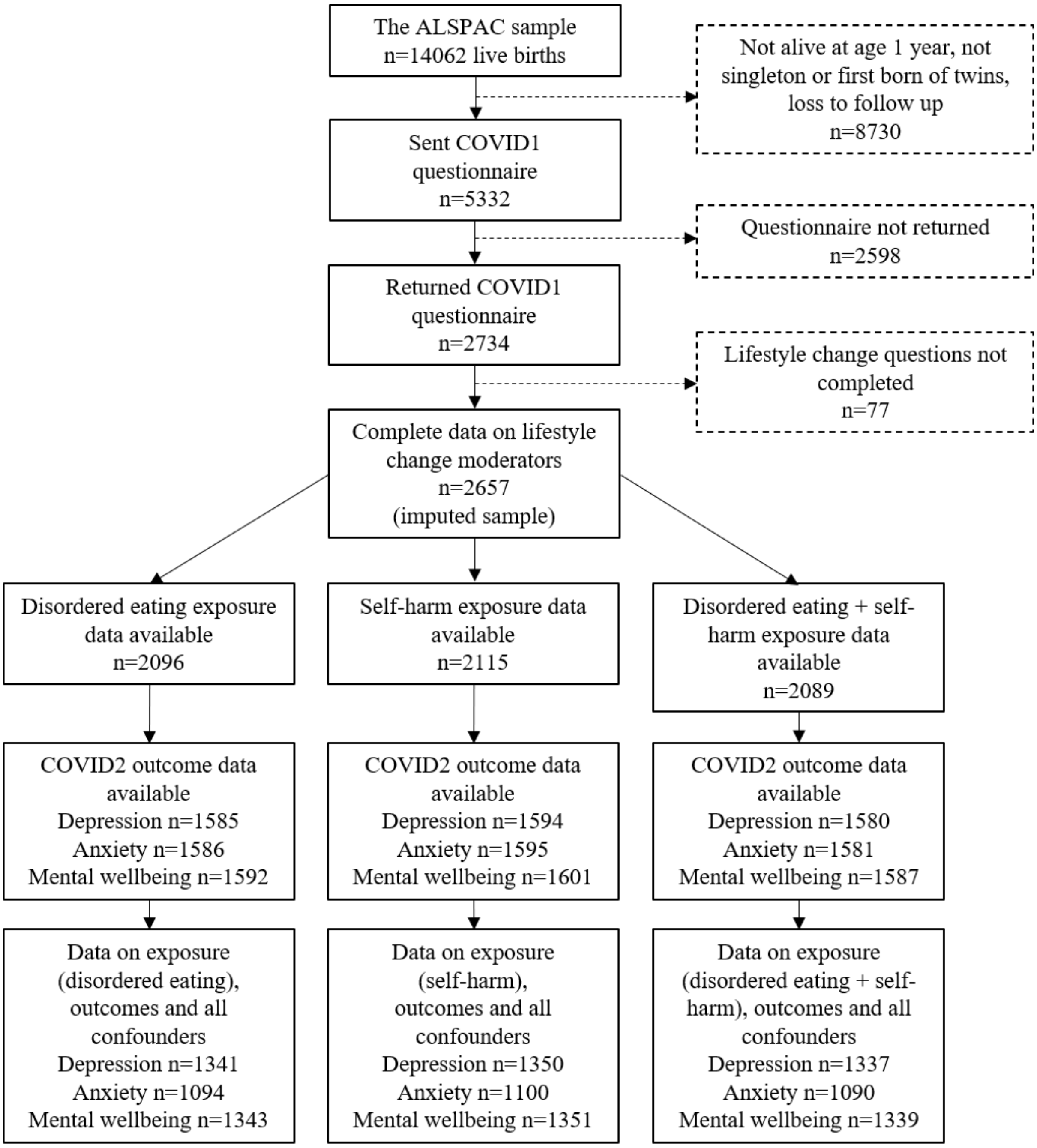
